# Impact of clonal hematopoiesis on atherosclerotic cardiovascular disease according to low-density lipoprotein cholesterol levels

**DOI:** 10.1101/2022.06.09.22276170

**Authors:** Heesun Lee, Han Song, Youngil Koh, Hyo Eun Park, Ji Won Yoon, Min Joo Kim, Soie Chung, Jung Ho Bae, Seung Ho Choi, Su-Yeon Choi, Bon-Kwon Koo

## Abstract

**Background:** Clonal hematopoiesis of indeterminate potential (CHIP), defined as a clonal expansion of hematopoietic stem cells with age-related recurrent somatic mutations, has recently emerged as a novel cardiovascular risk factor. However, the pathogenic mechanism of CHIP in atherosclerotic cardiovascular disease (ASCVD) development is not yet fully understood.

**Methods:** In a cohort comprising 4,300 asymptomatic Korean participants aged 40–79 years, we investigated the risks associated with ASCVD by CHIP and the interplay of CHIP with other traditional risk factors for developing ASCVD. Using coronary computed tomography angiography (CCTA), we noninvasively assessed the changes in coronary arteries to evaluate CHIP-associated atherosclerosis.

**Results:** CHIP was present in 368 participants (8.6%). The most commonly mutated genes in CHIP included *DNMT3A, TET2*, and *ASXL1*. During follow-up (median, 4.7 years), 18 ASCVD cases (4.9%) were observed in CHIP carriers vs. 62 (1.6%) in non-carriers (*p*<0.001). The presence of CHIP was associated with an increased risk of ASCVD (adjusted hazard ratio [HR] 2.54, 95% confidence interval [CI] 1.48–4.37, *p*<0.001) after adjusting for other conventional cardiovascular risk factors. Particularly, in participants with high levels of low-density lipoprotein (LDL) cholesterol, CHIP enhanced the risk of ASCVD development (adjusted HR 4.18, 95% CI 1.99–8.77, *p*<0.001), demonstrating a significant synergism between CHIP and LDL cholesterol (*S*-index, 4.61; 95% CI 1.04–20.57, *p*=0.045). Serial CCTAs supported our findings by displaying de novo measurable coronary atherosclerosis with unstable plaque and in proximal lesions only in CHIP carriers with high LDL cholesterol.

**Conclusion:** The presence of CHIP was significantly associated with the risk of ASCVD through synergy with LDL cholesterol in the general population, which might promote the early stage of coronary atherosclerosis.

## INTRODUCTION

Cardiovascular disease (CVD) has been the leading cause of death worldwide, and thus, cardiovascular risk factors have been evaluated to predict and prevent CVD.^1^ However, increasing evidence shows that these conventional risk factors are incompletely predictive of further CVD, particularly in elderly individuals, implying that there may still be unidentified risk factors in relation to aging.^2,3^ In a previous study including five community-based cohorts, lifetime risk estimates for CVD increased with aging, even for those with optimal risk factors during middle age.^2^ Hence, recent clinical efforts have been focused on discovering new cardiovascular risk factors.

Aging is associated with an increased frequency of somatic mutations in hematopoietic stem cells. Recent studies have revealed that clonal hematopoiesis of indeterminate potential (CHIP), defined as the clonal expansion of hematopoietic cells with aging-related recurrent somatic mutations in the absence of other hematologic abnormalities, has a close association with atherosclerotic CVD (ASCVD).^4,5^ Several experimental models have supported that hyperlipidemic mice with CHIP develop atherosclerosis due to inflammation promoted primarily via the interleukin (IL)-1β/IL-6 pathway.^5-9^ Furthermore, a study using a coronary artery calcium scan showed that coronary artery calcium scores (CACS) were higher in subjects with CHIP than in those without.^5^ Contrary to the prior results emphasizing the causal role of CHIP in ASCVD development, Heyde *et al*. proposed the conflicting hypothesis that CHIP could be a symptom rather than a cause of atherosclerosis, as atherosclerosis increased hematopoietic stem cell division, resulting in accelerated somatic evolution.^10^ Despite the recent accumulation of preclinical and clinical data regarding CHIP, the mechanism by which CHIP promotes the development of ASCVD is not fully understood, yet.

Here, we sought to investigate the interplay among CHIP, conventional risk factors, and ASCVD and to comprehensively analyze changes in coronary atherosclerosis using coronary computed tomography angiography (CCTA), in order to provide further information on CHIP-associated atherosclerosis in the general population.

## METHODS

### Study design and population

We collected blood samples from participants during a routine health check-up for screening purposes at the Seoul National University Hospital Healthcare System Gangnam Center between January 2014 and January 2017. Blood samples were then stored in a biorepository as a part of the Gene-Environmental Interaction and phEnotype (GENIE) cohort (n=10,348).^11^

From the GENIE cohort, we retrospectively recruited the study population who satisfied the inclusion criteria: those aged ≥65 years or those aged ≥40 years with more than one of the ASCVD risk factors (n=4,335). After excluding participants ≥80 years (n=13) and those who failed the quality control test for the blood samples (n=2) or did not have results regarding essential laboratory tests (n=20), 4,300 participants were finally analyzed (**Figure 1**). The ASCVD risk factors included hypertension, diabetes mellitus, dyslipidemia, chronic kidney disease, and current smoking. The ASCVD risk factors used in the criteria for study participant enrollment were defined as follows: hypertension as a systolic blood pressure (BP) ≥140 mmHg or diastolic BP ≥90 mmHg and/or the use of anti-hypertensive medications; diabetes mellitus as a fasting glucose ≥126 mg/dl or HbA1c ≥6.5%, and/or treatment by an oral hypoglycemic agent or insulin; dyslipidemia as total cholesterol ≥240 mg/dl, low-density lipoprotein (LDL) cholesterol ≥160 mg/dl, triglycerides ≥200 mg/dl, high-density lipoprotein (HDL) cholesterol <40 mg/dl, and/or the use of statin; chronic kidney disease as estimated glomerular filtration rate <60 ml/min/1.73 m^2^; current smoking as lifetime smoking ≥100 cigarettes and current consumption of at least 1 cigarette a day.

**Figure 1.**
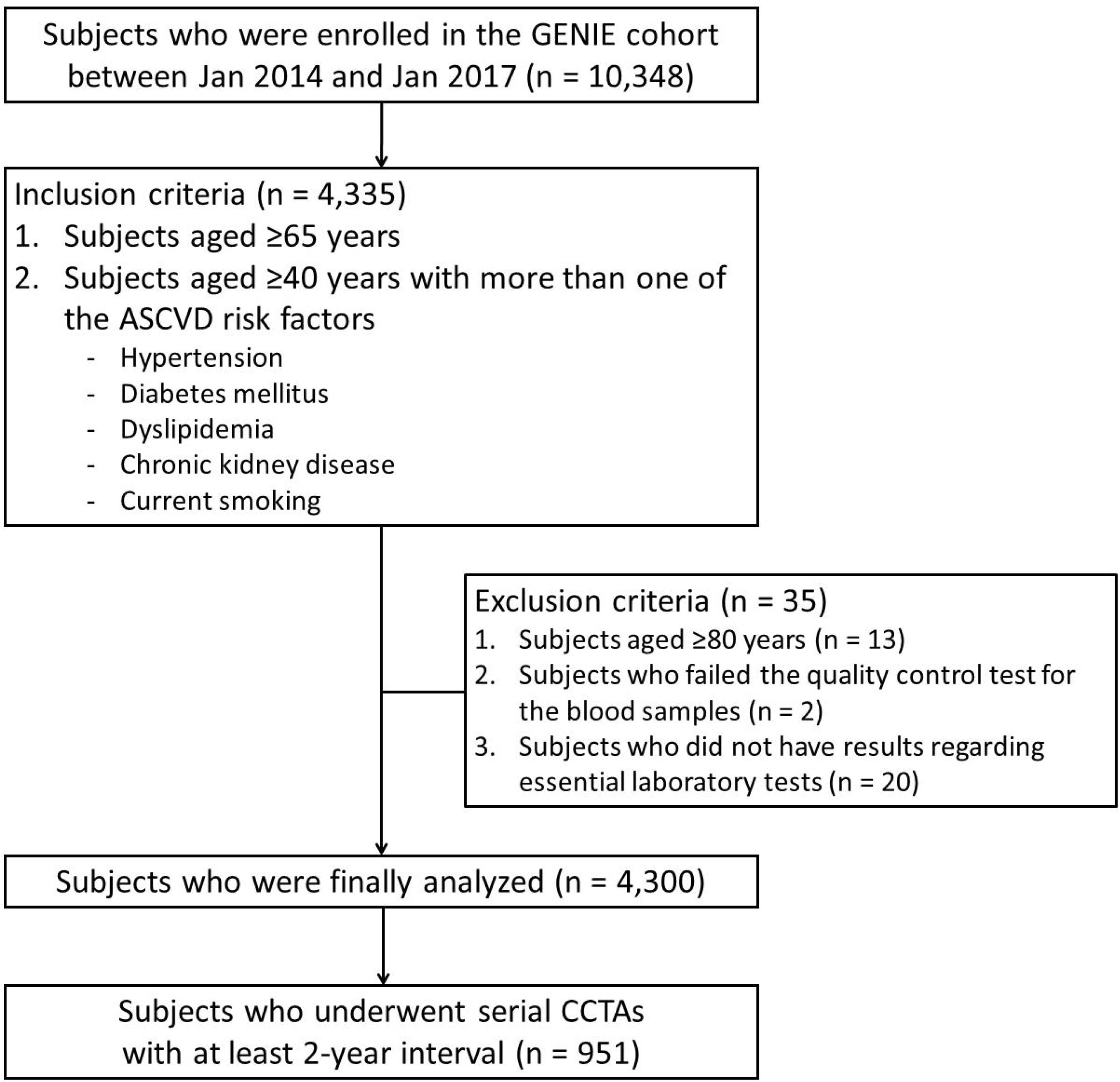
Schematic flow of the study population selected. ASCVD, atherosclerotic cardiovascular disease; CCTA, coronary computed tomography angiography; GENIE, Gene-ENvironmental Interaction and phEnotype; Jan, January.

The study protocol conformed to the ethical guidelines set by the Declaration of Helsinki. The institutional review board of our institution approved the GENIE cohort (H-1103-127-357), and this study (H-1908-121-1056). Informed consent for blood storage was obtained when collecting blood samples, and that for this study was waived owing to the retrospective nature.

### Targeted gene sequencing

Targeted gene sequencing was performed using the SureSelectXT HS custom panel (Agilent, Santa Clara, CA, USA) and NovaSeq 6000 (Illumina, San Diego, CA, USA) with 2×150 bp paired-end reads with a minimum coverage of 800x. The custom panel comprised a total exon of 89 genes frequently involved in CHIP (*APC, ASXL1, ASXL2, ATM, BCL11B, BCOR, BCORL1, BIRC3, BRAF, BRCC3, CARD11, CASP8, CBL, CD58, CD79B, CHEK2, CNOT3, CREBBP, CUX1, DDX3X, DNMT3A, EP300, ETV6, EZH2, FAM46C, FBXW7, FLT3, FOXP1, GNAS, GNB1, GPS2, HIST1H1C, IDH2, IKZF1, IKZF2, JAK1, JAK2, JAK3, JARID2, KDM6A, KIT, KLHL6, KMT2D, KRAS, LUC7L2, MAP3K1, MPL, MYD88, NF1, NFE2L2, NOTCH1, NOTCH2, NRAS, PDS5B, PDSS2, PHF6, PHIP, PIK3CA, PIK3R1, PPM1D, PRDM1, PRPF40B, PTEN, PTPN11, RAD21, RIT1, RPS15, SETD2, SETDB1, SF1, SF3A1, SF3B1, SMC1A, SMC3, SRSF2, STAG1, STAG2, STAT3, SUZ12, TBL1XR1, TET1, TET2, TNFAIP3, TNFRSF14, TP53, U2AF1, VHL, WT1, and ZRSR2*) (**Table S1**).^4,5,12-14^ The sequencing reads were aligned to the human genome (hg19) using BWA-MEM (v0.7.10) and GATK light (v2.3.9). Single-nucleotide variants (SNVs) were called using SNVer (v0.4.1), LoFreq (v0.6.1), and GATK UnifiedGenotyper (v2.3.9). Insertions and deletions were called using an in-house caller.^15^ Then, the unreliable variants that did not meet the following criteria were filtered as sequencing artifacts or germline variants: 1) number of altered reads at positive and negative strands ≥5 with mapping quality ≥30 and base quality ≥30; 2) variant allele frequency between 1.5–30%; 3) not among common germline variants including genomAD, 1k Genome v3, ESP6500, and ExAC; 4) not listed in the artifact database with minor allele frequency >2% in the internal panel of 1,000 Korean individuals.

### Variant annotation

All reliable non-synonymous oncogenic variants that had evidence of functional relevance in cancer were annotated as CHIP-driving mutations according to the following criteria: 1) any truncating mutations, including nonsense mutation, splice site mutation, or frameshift indel; 2) any variants previously reported as somatic on at least 20 occasions with solid cancer or on 10 occasions with hematopoietic and lymphoid cancer in the COSMIC database v83.

### CCTA image acquisition and analysis

To delineate the change in coronary arteries according to CHIP status, we collected the data of subjects who underwent baseline CCTA 2 years before and after the blood test for the GENIE cohort and follow-up CCTA with at least a 2-year interval (n=951) (**Figure 1**). CCTA images were acquired using either a retrospective electrocardiography (ECG)-gated or prospective ECG*-*triggered protocol by a 16- (SOMATOM Sensation 16; Siemens Medical Solutions, Forchheim, Germany) or 256-detector row CT scanner (Brilliance iCT 256; Philips Medical Systems, Cleveland, OH, USA), according to the Society of Cardiovascular Computed Tomography guidelines.^16^ All images were independently analyzed by two experienced radiologists who were blind to the clinical information. The CACS was calculated semi-quantitatively using the Agatston system (in units)^17^ with dedicated software (Rapidia 2.8; INFINITT, Seoul, Korea). The presence, extent, severity, location, and composition of coronary atherosclerotic plaques were assessed by per-segment and per-patient analyses with the modified 15-segment criteria.^18^ The presence of a coronary plaque was indicated by the detection of any clearly distinguishable lesion >1 mm^2^ within or adjacent to the coronary lumen in ≥2 image planes of ≥2 mm sized coronary arteries. The extent of coronary atherosclerosis was expressed using the total number of coronary artery segments and vessels with any plaque.^18^ The severity of focal maximal diameter stenosis was quantified by visual estimation, with an agreement between two independent radiologists. The plaque composition was categorized as noncalcified (<30% calcified plaque volume), mixed (30–70%), or calcified (>70%), with the calcified component being >130 Hounsfield units.^18^

The progression of coronary artery calcification was defined as the difference of ≥2.5 between the square roots of the baseline and follow-up CACS (Δ√transformed CACS).^19^ The changes in coronary atherosclerosis were explained by newly appeared or progressed plaques with measurable diameter stenosis, defined as luminal narrowing of >30%, at the corresponding segment in the follow-up image.^20^

### Study outcome

The study outcome was on ASCVD, and the outcome data were collected up to March 2020 using hospital admissions confirmed from electronic medical records with one or more relevant International Classification of Disease-10 codes, ascertainment of mortality determined via the Ministry of Security and Public Administration, and by self-report from participants. ASCVD was defined as a composite of cardiovascular death, acute myocardial infarction, coronary revascularization, definite angina not followed by coronary revascularization, stroke, carotid artery disease with revascularization, aortic aneurysm with percutaneous endovascular stent graft repair or surgical repair, and peripheral artery disease (PAD) with percutaneous angioplasty, open revascularization or amputation.^21^ Death was considered to be related to a cardiovascular cause if it occurred from coronary heart disease (I20 – I25) or stroke (I61, I63, or I64). For acute myocardial infarction, one or more relevant ICD-10 codes (I21 – I23) at discharge were included. Revascularization included cases with coronary artery bypass grafting or coronary angioplasty with or without stenting. Definite angina was defined as typical chest pain with objective evidence of reversible myocardial ischemia or obstructive coronary artery disease on invasive coronary angiography. Stroke was based on the rapid onset of a documented focal neurologic deficit lasting 24 hours with accompanying evidence of a clinically relevant lesion on brain imaging with the diagnosis codes of I61, I63, or I64, and no nonvascular cause was identified and excluded. Carotid artery disease was defined as significant carotid stenosis with percutaneous endovascular stent graft repair or surgical repair of the carotid artery under the diagnosis code of I65.2, and aortic disease was defined as aortic aneurysm with percutaneous endovascular stent graft repair or surgical repair of the aorta under the diagnosis codes of I70 and I71. For PAD, any percutaneous intervention or surgery for peripheral arteries under the diagnosis codes of I70.2, I70.9, I73.1, or I73.8-9 was included.

### Statistical analysis

Continuous variables were presented as the mean ± standard deviation or median with interquartile range and were compared using Student’s t test for independent samples or the Mann–Whitney test. Categorical variables were expressed as numbers and percentages and compared using the χ^2^ test or Fisher’s exact test. The associations of the presence of CHIP with incident ASCVD and CCTA outcomes were tested using a Cox proportional hazard model with covariates including age, sex, and other related risk factors. Event-free survival from incident ASCVD was estimated using the Kaplan–Meier method and compared between the groups by the log-rank test. The relative excess risk from synergism of CHIP and high LDL cholesterol levels was calculated from the synergy index (*S*) using the R epiR package (R Foundation for Statistical Computing, Vienna, Austria). For this analysis, participants were included in the high LDL cholesterol group if their LDL cholesterol level was ≥160 mg/dl (classified as ‘high level’ by the National Cholesterol Education Program^22,23^) or if they were undergoing statin therapy, and those without measured LDL cholesterol levels were excluded. Two-sided *p* values <0.05 were considered statistically significant. All statistical analyses were performed using Python version 3.7.9 (Python Software Foundation) and its packages Numpy (1.19.4), Scipy (1.5.3), Scikit-learn (0.23.2), and Lifelines (0.25.6).

## RESULTS

### Baseline characteristics of the study population

Among the 4,300 participants (mean age, 55.7 years; males, 74.1%), CHIP was identified in 368 (8.6%). Detailed clinical characteristics according to the CHIP status are shown in **Table 1**. The most commonly mutated genes in CHIP included *DNMT3A, TET2*, and *ASXL1*, in order (**Figure 2A, Table S1**). The prevalence and size of CHIP increased with age (**Figure 2B**). However, there was no sex difference in the size of CHIP clones (4.9% in males vs. 5.0% in females; *p*=0.916).

**Table 1.** Clinical characteristics of the study population according to CHIP status.

|  | **Total**<br>**(n = 4,300)** | **CHIP carrier**<br>**(n = 368)** | **Non-carrier**<br>**(n = 3,932)** | **P** |
| --- | --- | --- | --- | --- |
| Age | 55.7±8.5 | 60.3±8.6 | 55.3±8.3 | <0.001 |
| Follow up duration, years | 4.7 (3.2-5.3) | 4.9 (3.3-5.3) | 4.7 (3.1-5.3) | 0.655 |
| Male | 3,185 (74.1) | 259 (70.4) | 2,926 (74.4) | 0.093 |
| Smoking | 2,635 (61.3) | 222 (60.3) | 2,413 (61.4) | 0.696 |
| Current smoking | 1,184 (27.5) | 77 (20.9) | 1,107 (28.2) | 0.003 |
| Former smoking | 1,451 (33.7) | 145 (39.4) | 1,306 (33.2) | 0.018 |
| Body mass index, kg/m<sup>2</sup> | 24.2±2.8 | 24.1±2.4 | 24.2±2.8 | 0.275 |
| BMI≥25 kg/m<sup>2</sup> | 1,591 (37.0) | 130 (35.3) | 1,461 (37.2) | 0.499 |
| Systolic BP, mmHg | 119.5±13.3 | 120.6±13.3 | 119.3±13.3 | 0.084 |
| Diastolic BP, mmHg | 79.3±10.0 | 78.3±8.8 | 79.4±10.1 | 0.042 |
| Hypertension | 1,811 (42.1) | 154 (41.8) | 1,657 (42.1) | 0.956 |
| Anti-hypertensive medication | 1,067 (24.8) | 109 (29.6) | 958 (24.4) | 0.027 |
| Diabetes mellitus | 691 (16.1) | 62 (16.8) | 629 (16.0) | 0.656 |
| Anti-diabetic medication | 265 (6.2) | 22 (6.0) | 243 (6.2) | 1.000 |
| Dyslipidemia | 2,561 (59.6) | 212 (57.6) | 2,349 (59.7) | 0.437 |
| Lipid-lowering medication | 745 (17.3) | 68 (18.5) | 677 (17.2) | 0.564 |
| Total cholesterol, mg/dl | 195.6±39.1 | 190.8±37.0 | 196.1±39.2 | 0.012 |
| LDL cholesterol, mg/dl | 124.5±33.7 | 120.2±31.4 | 124.9±33.9 | 0.012 |
| HDL cholesterol, mg/dl | 51.2±12.2 | 51.7±12.9 | 51.1±12.2 | 0.374 |
| Triglycerides, mg/dl | 109.0<br>(77.0-160.0) | 103.0<br>(74.0-153.0) | 110.0<br>(77.2-160.0) | 0.056 |
| Fasting glucose, mg/dl | 104.7±21.6 | 105.4±21.4 | 104.7±21.6 | 0.529 |

2 Values are the mean ± standard deviation, median (interquartile range), or n (%). BMI, body 3 mass index; BP, blood pressure; CHIP, clonal hematopoiesis of indeterminate potential; HDL, 4 high-density lipoprotein; LDL, low-density lipoprotein.

**Figure 2.**
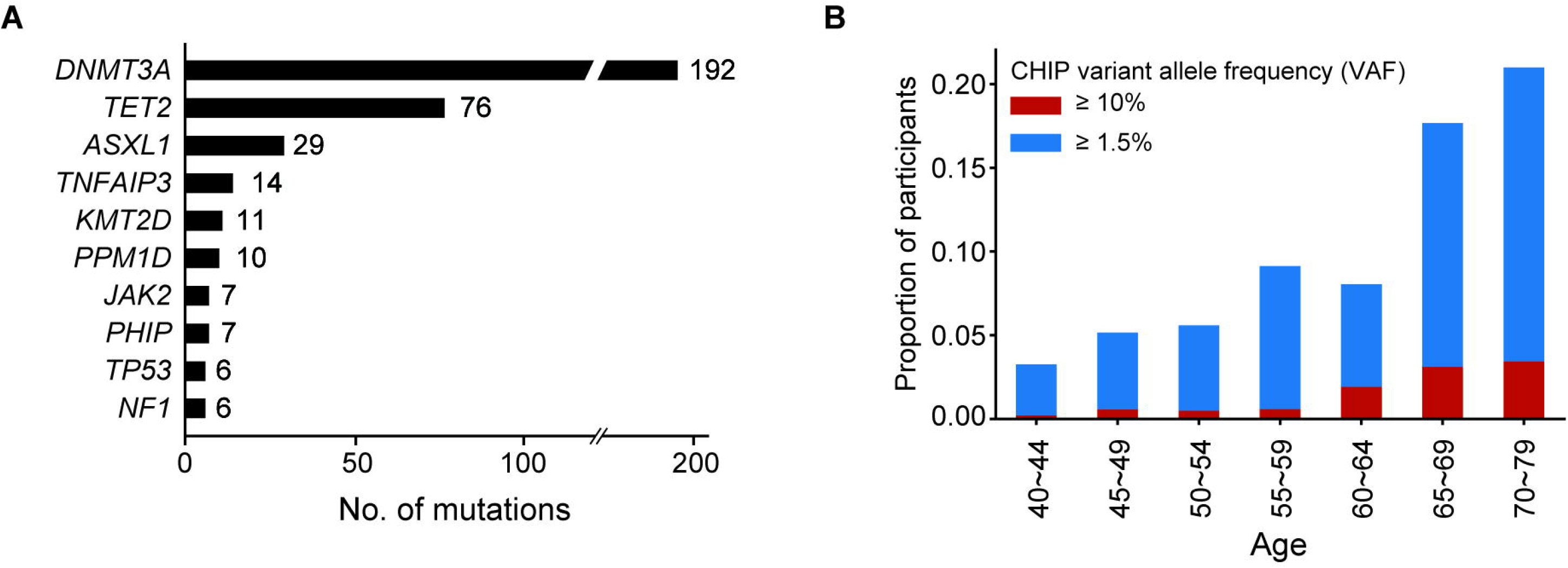
Baseline genetic characteristics in the study population. (A) The ten most common genes harboring somatic mutations associated with CHIP were drawn. In the present cohort, somatic mutations in the *DNMT3A* gene were the most frequent (46.7%). (B) The prevalence and clone size of CHIP increased with age. CHIP, clonal hematopoiesis of indeterminate potential; No, number.

### Incidence and risk factors for ASCVD

During follow-up (median, 4.7 years), ASCVD occurred in 80 participants (1.9%), including 1 cardiovascular death, 2 acute myocardial infarction, and 56 revascularizations (**Table S2**). The incidence of ASCVD was higher in CHIP carriers than in non-carriers (18 [4.9%] vs. 62 [1.6%]; *p*<0.001). The Kaplan–Meier curves also demonstrated that CHIP carriers had worse prognosis than non-carriers (log-rank *p*<0.001) (**Figure 3A**). In the Cox regression analysis (**Table 2**), conventional risk factors, including old age, male sex, hypertension, diabetes mellitus, and dyslipidemia, were independently associated with the risk of ASCVD. The risk of ASCVD was significantly increased when an individual had CHIP (hazard ratio [HR], 2.54; 95% confidence interval [CI], 1.48–4.37; *p*<0.001).

**Table 2.** The risk of clinical events by multivariable Cox regression analysis.

|  | Adjusted HR* (95% CI) | *p* |
| --- | --- | --- |
| Male | 6.55 (2.36–18.18) | <0.001 |
| Age (per 1 year increment) | 1.06 (1.03–1.09) | <0.001 |
| Obesity (BMI $\geq 25$ kg/m<sup>2</sup>) | 0.99 (0.63–1.56) | 0.955 |
| CHIP mutation | 2.54 (1.48–4.37) | <0.001 |
| Hypertension | 1.70 (1.07–2.70) | 0.024 |
| Diabetes mellitus | 2.37 (1.49–3.78) | <0.001 |
| Dyslipidemia | 1.72 (1.07–2.75) | 0.025 |
| Current smoking | 1.53 (0.91–2.56) | 0.110 |

\*Multivariable Cox regression model was adjusted for age, sex, hypertension, diabetes mellitus, dyslipidemia, obesity and current smoking status. CI, confidence interval; HR, hazard ratio; other abbreviations as **Table 1**.

**Figure 3.**
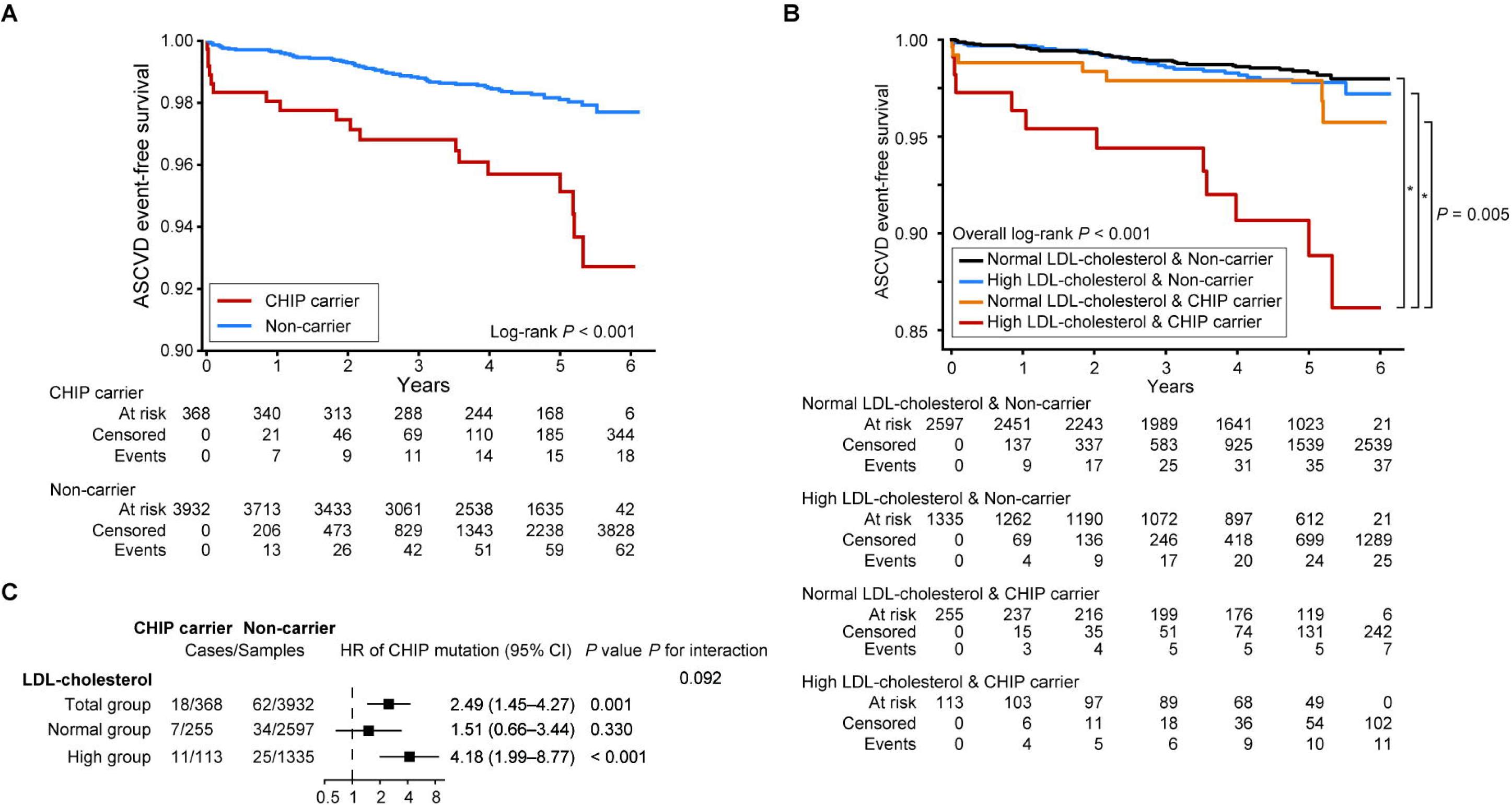
Association between CHIP and ASCVD and synergy between CHIP and LDL cholesterol levels for developing ASCVD. (A) The Kaplan–Meier curves for event-free survival showed that CHIP carriers had a worse prognosis during follow-up than non-carriers. (B) A comparison of the prognosis according to the presence of CHIP and LDL cholesterol levels revealed that CHIP carriers with high LDL cholesterol had the worst prognosis. * indicates *p*<0.001. (C) The risk of ASCVD increased by approximately 2.5 times in CHIP carriers. The increased risk of ASCVD by CHIP intensified under high LDL cholesterol (age- and sex-adjusted HR, 4.18; *p*<0.001), whereas no significant increase in risk was found under conditions of normal LDL cholesterol levels (age- and sex-adjusted HR, 1.51; *p*=0.330). CI, confidence interval; HR, hazard ratio; LDL, low-density lipoprotein; other abbreviations as **Figures 1 and 2**.

Among CHIP carriers, incident ASCVD was detected in 10 of 180 participants with *DNMT3A* mutations, 3 of 74 with *TET2* mutations, 2 of 8 with *PPM1D* mutations, and 4 of 124 with mutations in others. One participant had 2 mutations in *PPM1D* and *ASXL2* simultaneously. When analyzing the risk of ASCVD by mutations in *DNMT3A, TET2, ASXL1*, and *JAK2*, only *DNMT3A* mutation was associated with an increased risk of ASCVD (HR, 2.82; 95% CI, 1.44–5.51; *p*=0.002) (**Figure S1**).

### Impact of CHIP and cardiovascular risk factor coexistence

We further analyzed the risk of ASCVD development when CHIP coexisted with other cardiovascular risk factors to understand the interaction between genetic and clinical risk factors (**Table S3**). The risk of ASCVD increased with a stepwise pattern, depending on the combination of CHIP and LDL cholesterol (**Table 3**). In cases with either the presence of CHIP or high LDL cholesterol, the risk of ASCVD increased by approximately 50% in both groups. The risk of ASCVD was approximately six times higher in participants with CHIP and high LDL cholesterol than in those with no mutation and normal LDL cholesterol (HR, 5.98; *p*<0.001). Furthermore, CHIP and high LDL cholesterol demonstrated a significant synergism on the development of ASCVD (*S*-index, 4.61; *p*=0.045) (**Table 3**), but high total cholesterol or triglycerides or low HDL cholesterol did not. The Kaplan–Meier curves also showed that participants with high LDL cholesterol and CHIP had the worst prognosis (overall log-rank, *p*<0.001). However, no statistically significant difference in survival analysis was observed among groups other than the group with CHIP and high LDL cholesterol (**Figure 3B**).

**Table 3.** Synergism analysis of CHIP status and LDL cholesterol levels.

|  | ASCVD events<br>(n = 80) | No event<br>(n = 4,220) | Adjusted HR<br>(95% CI)* | p | Synergy index<br>(95% CI)<sup>†</sup> | p |
| --- | --- | --- | --- | --- | --- | --- |
| *LDL-c levels and CHIP status* |  |  |  |  |  |  |
| Normal LDL-c and non-carrier | 37 (46.3) | 2,560 (60.7) | 1.00<br>(reference) | - |  |  |
| High LDL-c and non-carrier | 25 (31.3) | 1,310 (31.0) | 1.52 (0.91–2.53) | 0.106 |  |  |
| Normal LDL-c and CHIP carrier | 7 (8.8) | 248 (5.9) | 1.56 (0.69–3.53) | 0.287 |  |  |
| High LDL-c and CHIP carrier | 11 (13.8) | 102 (2.4) | 5.98 (3.03–11.80) | <0.001 | 4.61 (1.04–20.57) | 0.045 |

\*The risk of ASCVD events was adjusted for age and sex. <sup>†</sup>Synergy index (S-index) was calculated from $(RR_{A+B+}-1)/((RR_{A+B-}-1)+(RR_{A-B+}-1))$ .
3 $RR_{AB}$ is the relative risk of disease if both factors A and B are present or absent. ASCVD, atherosclerotic cardiovascular disease; LDL-c, low- 4 density lipoprotein cholesterol; other abbreviations as **Tables 1 and 2**.

Similar to the synergism analysis, high LDL cholesterol intensified the risk of ASCVD by CHIP (HR, 4.18; *p*<0.001), while normal LDL cholesterol attenuated it (HR, 1.51; *p*=0.330) (**Figure 3C**). It also highlighted the synergistic effect between CHIP and LDL cholesterol, given that the clone sizes of both groups with high and normal LDL cholesterol were similar (**Figure S3**).

Sensitivity analysis after adjusting for diabetes mellitus and smoking, which showed a significant association with CHIP in previous studies^24,25^, was performed to demonstrate the direct association between CHIP and LDL cholesterol. Likewise, a synergistically increased risk of ASCVD in CHIP carriers with high LDL cholesterol was found. (**Table 4**).

**Table 4.** Sensitivity analysis for synergism between CHIP status and LDL cholesterol levels.

|  | ASCVD events<br>(n = 80) | No event<br>(n = 4,220) | Adjusted HR<br>(95% CI)* | *p* | Synergy index<br>(95% CI)<sup>†</sup> | *p* |
| --- | --- | --- | --- | --- | --- | --- |
| *LDL-c levels and CHIP status* |  |  |  |  |  |  |
| Normal LDL-c and non-carrier | 37 (46.3) | 2,560 (60.7) | 1.00<br>(reference) | - |  |  |
| High LDL-c and non-carrier | 25 (31.3) | 1,310 (31.0) | 1.60 (0.96–2.67) | 0.070 |  |  |
| Normal LDL-c and CHIP carrier | 7 (8.8) | 248 (5.9) | 1.62 (0.71–3.66) | 0.251 |  |  |
| High LDL-c and CHIP carrier | 11 (13.8) | 102 (2.4) | 6.32 (3.19–12.52) | <0.001 | 4.37 (1.08–17.64) | 0.038 |

\*The risk of ASCVD events was adjusted for age, sex, diabetes and current smoking status, which showed a significant association with CHIP in previous studies. <sup>†</sup>Synergy index (S-index) was calculated from $(RR_{A+B+}-1)/((RR_{A+B-}-1)+(RR_{A-B+}-1))$ . $RR_{AB}$ is the relative risk of disease if both factors A and B are present or absent. Abbreviations as **Tables 1, 2, and 3**.

The presence of CHIP provided additional risk stratification for ASCVD when concomitant with hypertension, diabetes mellitus, smoking, or obesity (**Figure S2**), but there was no significant synergy between CHIP and these clinical risk factors. The coexistence of CHIP with diabetes mellitus tended to synergistically increase the risk of ASCVD but failed to show statistical significance (*S*-index, 3.00; *p*=0.069).

### Changes in coronary atherosclerosis by CHIP according to LDL cholesterol

Among 951 participants who underwent serial CCTAs (median interval, 5.4 years), 74 (7.8%) had CHIP. Although the mean age at which baseline CCTA was performed was higher in CHIP carriers than in non-carriers, no remarkable differences in baseline CCTA findings were demonstrated, except for the severity of maximal stenosis (**Table S4**). Furthermore, no remarkable differences in the changes at follow-up CCTA were noted, except that coronary atherosclerosis in the proximal lesions was likely to deteriorate in CHIP carriers (17.6% vs. 8.4%; HR, 1.72; *p*=0.081) (**Figure 4A**). However, it is noteworthy that the changes in coronary atherosclerosis on CCTA differed according to LDL cholesterol. In participants with high LDL cholesterol (n=287), CHIP led to the development of de novo measurable coronary atherosclerosis, mainly composed of mixed or noncalcified plaques, particularly at crucial locations, including the left main or proximal left anterior descending coronary arteries (**Figure 4B**). In contrast, in participants with normal LDL cholesterol (n=664), the pattern of progression in coronary atherosclerosis did not change depending on whether CHIP was present (**Figure 4C**).

**Figure 4.**
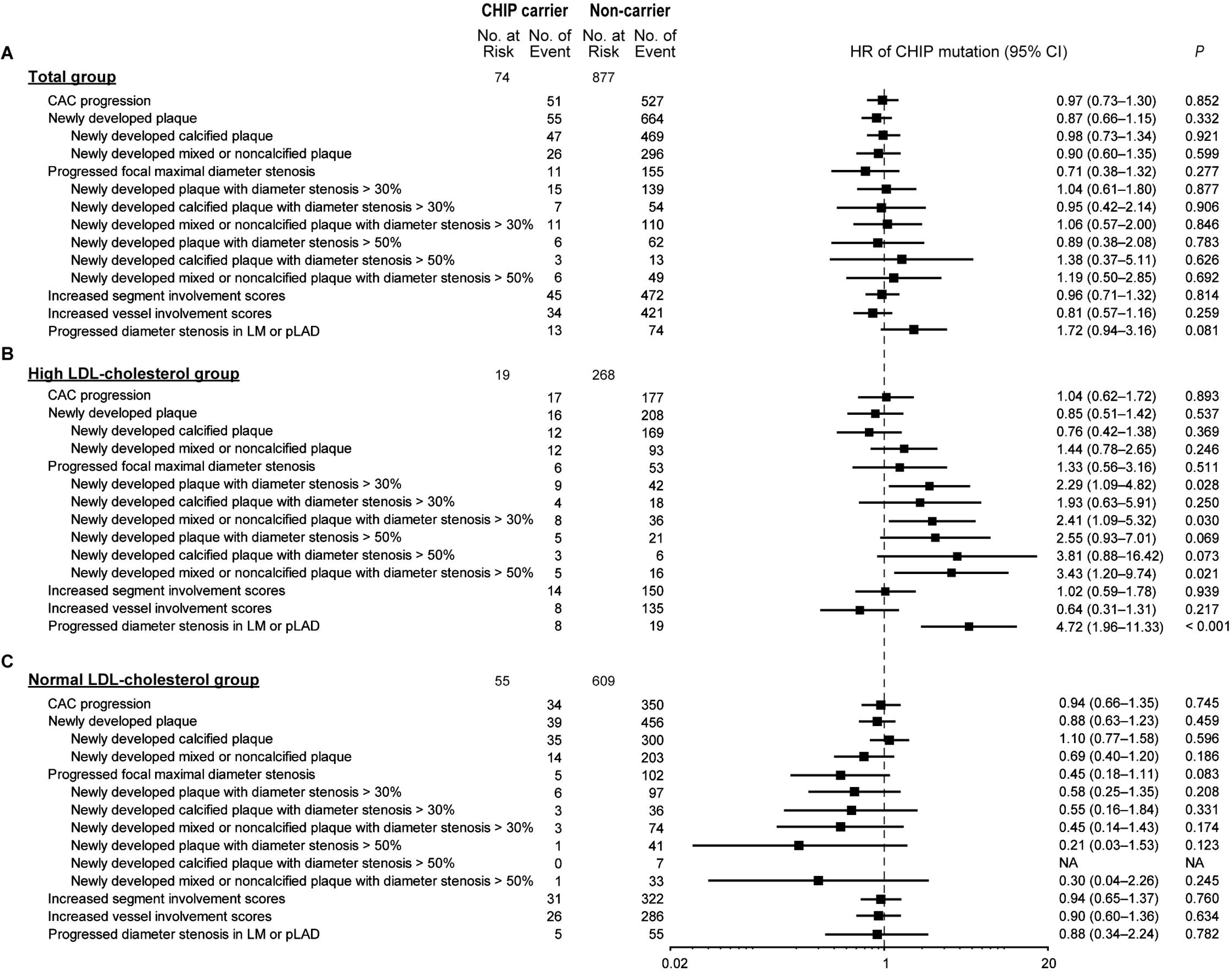
Changes in coronary atherosclerosis by CHIP according to LDL cholesterol levels. (A) There was no significant difference in the changes in the coronary tree between CHIP carriers and non-carriers in the total cohort, except the progression of coronary atherosclerosis in the proximal lesions. However, the changes in coronary atherosclerosis on CCTA differed according to LDL cholesterol levels. (B) In participants with high LDL cholesterol, new measurable unstable plaques, mainly in the proximal lesion, were developed by CHIP, (C) whereas the pattern of progression in coronary atherosclerosis did not change depending on the CHIP status in those with normal LDL cholesterol levels. All models were adjusted by age and sex. CAC, coronary artery calcification; LM, left main; pLAD, proximal left anterior descending coronary artery; CCTA, coronary computed tomography angiography; other abbreviations as **Figures 1, 2, and 3**.

## DISCUSSION

The main findings of this study consisting of 4,300 community-dwelling asymptomatic adults aged 40–79 years were as follows: 1) the presence of CHIP (368/4,300; 8.6%) was significantly associated with the risk of ASCVD, with an HR of 2.54, even after adjusting for other cardiovascular risk factors; 2) the risk of ASCVD by CHIP was intensified when coexistent with high LDL cholesterol, suggesting a synergism between CHIP and LDL cholesterol; and 3) serial CCTAs supported our findings by demonstrating that CHIP-associated coronary atherosclerosis was characterized by new measurable unstable plaques in the proximal lesion only under high LDL cholesterol.

### Association between CHIP and ASCVD

Since the first study reporting that age-related CHIP increased the risk of mortality, mainly due to the risk of ASCVD,^4^ several studies have constantly demonstrated a close association of CHIP with atherosclerosis and subsequent ASCVD. In a case–control study by Jaiswal *et al*., the presence of CHIP was associated with an increased risk of CHD and early-onset myocardial infarction, which was supported by findings of accelerated atherosclerosis in mutated mice and higher CACS in CHIP carriers.^5^ Similarly, in a study using the UK Biobank data, CHIP had a significant association with the increased risk of ASCVD with proportion to the clone sizes.^9^ Several research groups have attempted to explain how CHIP promotes ASCVD based on preclinical results in murine models, emphasizing the importance of proinflammatory cytokines, including IL-1β and IL-6.^5-8^ Consistent with these prior results, the present study verified the close association between CHIP and ASCVD in an Asian general population. In the current study, CHIP was observed in 8.6% of the study population, and the incidence rate of ASCVD was more than three times higher in CHIP carriers. The risk of ASCVD by CHIP independently increased by approximately 2.5 times, which was comparable with that by diabetes mellitus. In the analysis according to each gene, the *DNMT3A* mutation, one of the most commonly noted genes related to ASCVD in previous studies,^5,9,31^ showed a significant association with the risk of ASCVD.

### Reciprocity between CHIP and conventional risk factors on the development of ASCVD

One of the main findings of this study is the identification of the synergy between CHIP and LDL cholesterol levels on ASCVD development. Dyslipidemia, particularly high LDL cholesterol, is well known to be associated with atherosclerosis and subsequent ASCVD.^26^ Therefore, many prior experiments have shown the contribution of CHIP driver genes toward atherosclerosis in animal models with knockout of the *Ldlr* gene or feeding a high fat/high cholesterol diet to establish the atherogenic environment and explore a causal link from CHIP to atherosclerosis.^5-8^ Interestingly, a recent study by Heyde *et al*.^10^ suggested an intricate interplay among chronically altered lipid level after atherogenic diet, promoted chronic vascular inflammation, and plaque formation, which was referred to as the atherosclerosis trait complex. In *Ldlr-*deficient mice with atherosclerosis trait complexes, hematopoietic stem cell proliferation was accelerated, and finally, the mutant fraction growth rate was significantly increased. It implicated a bidirectional relationship between CHIP and atherosclerosis by collaborating with LDL cholesterol levels. Considering this, we also disclosed synergism for the risk of ASCVD between CHIP and high LDL cholesterol. Moreover, we demonstrated that changes in coronary atherosclerosis were observed only in CHIP carriers with high LDL cholesterol but not in those with normal LDL cholesterol. It is evident that the disparate response of changes in coronary atherosclerosis depending on LDL cholesterol levels can explain our novel finding, in which the increased risk of ASCVD development by CHIP was more obvious under conditions of high LDL cholesterol levels, providing synergism for poor prognosis. As the first suggestion of the reciprocity between CHIP and LDL cholesterol on ASCVD development, we think that our results might provide a better understanding of CHIP-associated atherosclerosis in humans.

Metabolic factors other than LDL cholesterol may affect the impact of CHIP on ASCVD. We observed a tendency for synergy between CHIP and diabetes mellitus in ASCVD development. Although not statistically proven in our study, high glucose levels might be another reciprocal mechanism exaggerating the adverse effect of CHIP on ASCVD, as supported by a recent animal study in which *TET2* deficiency-driven CHIP aggravated insulin resistance in aged and obese mice.^25^ Further human studies focusing on the relationship between glucose levels and CHIP are required.

### Plausible mechanism of CHIP-associated atherosclerosis

The correlation between CHIP and the initiation/progression of atherosclerosis has been shown in various *in vitro* and *in vivo* studies.^5-8^ Among them, one study investigated the association of CHIP with atherosclerosis and ASCVD by employing CACS as a good predictor for ASCVD.^5^ However, since CACS is limited in recognizing specific changes in coronary arteries,^15^ we utilized CCTA to describe the changes of a coronary tree in detail^27^ and to provide accurate and intuitive information regarding atherosclerosis.^28^ In the current study, it was found that coronary atherosclerosis significantly deteriorated in CHIP carriers with high LDL cholesterol. Notably, it tended to cause new measurable coronary atherosclerosis in crucial lesions, such as the left main or proximal left anterior descending coronary arteries, mainly composed of lipid-rich plaques rather than calcified plaques. Given that atherosclerosis occurs predominantly in the proximal vessels and plenty of lipids inside indicates a fresh plaque,^29^ it can be assumed that early plaque formation following inflammation, especially LDL cholesterol-driven, might be a key mechanism that could explain the triangular relationship among CHIP, LDL cholesterol, and ASCVD. This was in line with a prior study where *Ldlr*^*-/-*^ mice with *JAK2*^*V617F*^ mutation, one of the most frequently mutated in CHIP, promoted early plaque formation and increased intralesional complexity.^7^ Additionally, macrophages are known to drive atherosclerosis along with cholesterol. CHIP, especially with *TET2*^30^ or *DNMT3A*^31^ mutations, renders macrophages more inflammatory. This inflammatory feature of macrophages in atherosclerotic plaques with CHIP might play an important role in plaque progression.

### Limitations

There are several limitations in this study. First, the timing of the blood test and CCTA were not identical, and the changes in CHIP status were not traced during the follow-up. However, considering that an independent association between CHIP and ASCVD remained in multivariable analysis and clearly differentiated results of follow-up CCTA were shown in CHIP carriers, concerns can be alleviated. Second, therapeutic procedures after the blood test were not guided by the prescribed protocols due to the retrospective nature of the study, and the clinical events might have been influenced by facultative medical decisions during follow-up. Last, the effect of each CHIP driver gene or the comparison according to clone sizes could not be assessed because of the relatively low-risk population and the limited number of clinical events. Further prospective studies including individuals with diverse risk profiles are warranted.

## CONCLUSIONS

The risk of ASCVD by CHIP synergistically increased with high LDL cholesterol levels but not with other lipid variables or other conventional cardiovascular risk factors. This finding was supported by the specific changes in coronary atherosclerosis on serial CCTAs, such as de novo measurable coronary atherosclerosis, mixed or noncalcified plaques and involvement of proximal lesions, all of which implied an early stage of atherosclerosis. Our findings might provide new insights for understanding CHIP-associated atherosclerosis, obtain persuasive evidence for the synergism between CHIP and LDL cholesterol to develop ASCVD, and potentially help manage further ASCVD in the general population.

## Data Availability

All data produced in the present study are available upon reasonable request to the authors

## ACKNOWLEDGEMENTS

The authors would like to thank Dr. Hogune Im and Dr. Choonghyun Sun for helpful assistance in next-generation sequencing and CHIP variant analysis.

