## Supplementary Figure Legend for "Impact of clonal hematopoiesis on atherosclerotic cardiovascular disease according to low-density lipoprotein cholesterol levels"

**SUPPLEMENTARY FIGURE LEGENDS**

**Figure S1. The risk of clinical events according to driver genes of CHIP.**

In the current cohort, the presence of CHIP increased the risk of ASCVD by approximately 2.5 times (HR, 2.49; *p=*0.001). When analyzed the risk of ASCVD by mutations in *DNMT3A*, *TET2*, *ASXL1* and *JAK2*, only *DNMT3A* mutation had an increased risk of ASCVD (HR, 2.82; *p*=0.002). Model was adjusted for age and sex. ASCVD, atherosclerotic cardiovascular disease; *ASXL1*, Additional sex combs-like 1; CHIP, clonal hematopoiesis of indeterminate potential; CI, confidence interval; *DNMT3A*, DNA methyltransferase 3 alpha; HR, hazard ratio; *JAK2*, Janus Tyrosine Kinase 2; No, number; *TET2*, Tet methylcytosine dioxygenase 2.

**Figure S2. Kaplan-Meier curves for event-free survival according to CHIP and clinical risk factors**

The presence of CHIP provided further risk stratification for ASCVD when concomitant with (A) hypertension, (B) diabetes mellitus, (C) smoking, or (D) obesity, suggesting the impact of the presence of CHIP on ASCVD as much as other conventional risk factors. DM, diabetes mellitus; HTN, hypertension; other abbreviations as **Figure S1**.

**Figure S3. Comparison of CHIP clone size according to LDL cholesterol levels**

The clone sizes between groups with high and normal LDL cholesterol levels were similar (*p* = 0.991). LDL, low-density lipoprotein; other abbreviations as **Figure S1**.
