## Supplementary figures and images for "Impact of clonal hematopoiesis on atherosclerotic cardiovascular disease according to low-density lipoprotein cholesterol levels"

### Supplementary figure 1

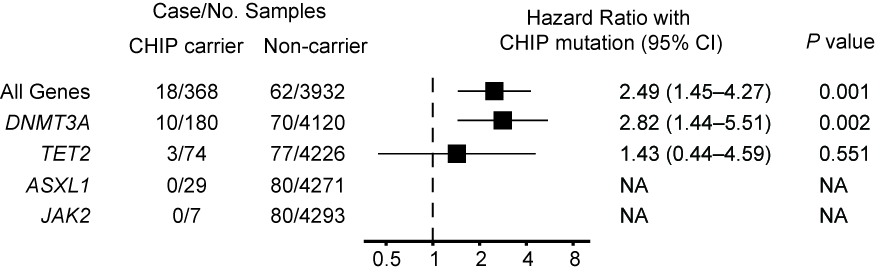

### Supplementary figure 2

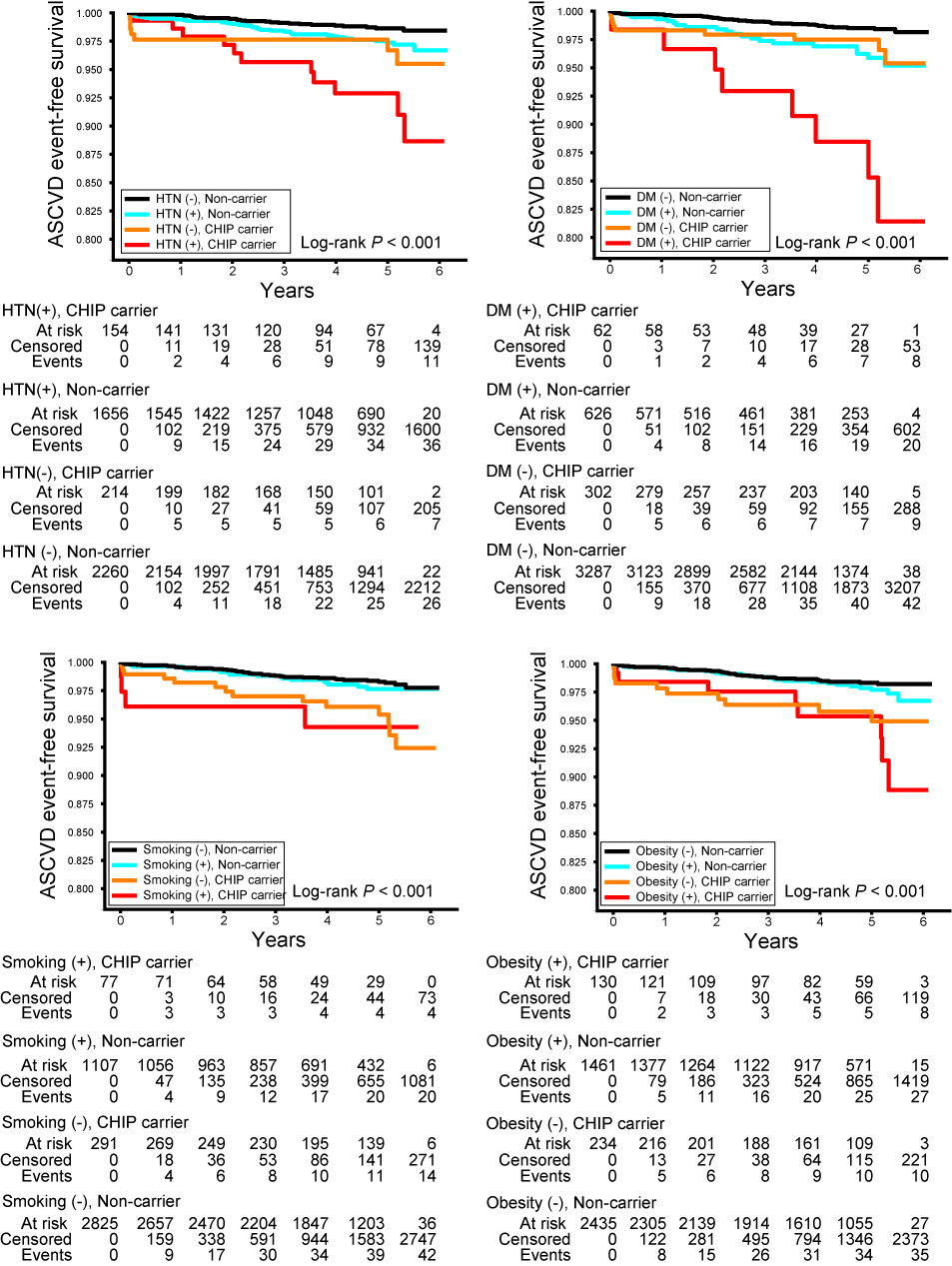

### Supplementary figure 3

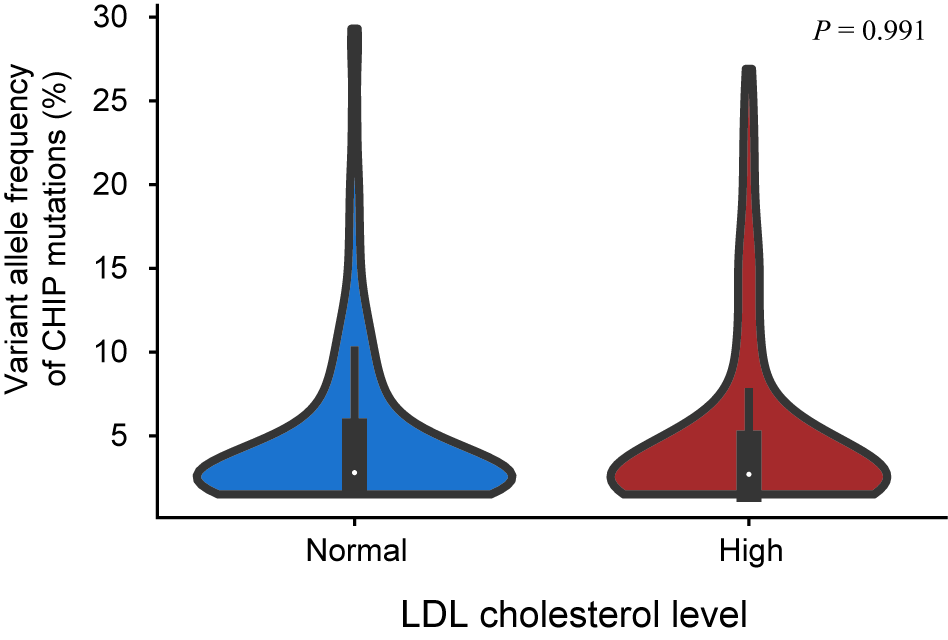
