## Supplementary Tables for "Impact of clonal hematopoiesis on atherosclerotic cardiovascular disease according to low-density lipoprotein cholesterol levels"

**Table S1. List of detected CHIP mutation in our study cohort**

(Attached with CHIP_list.pdf)

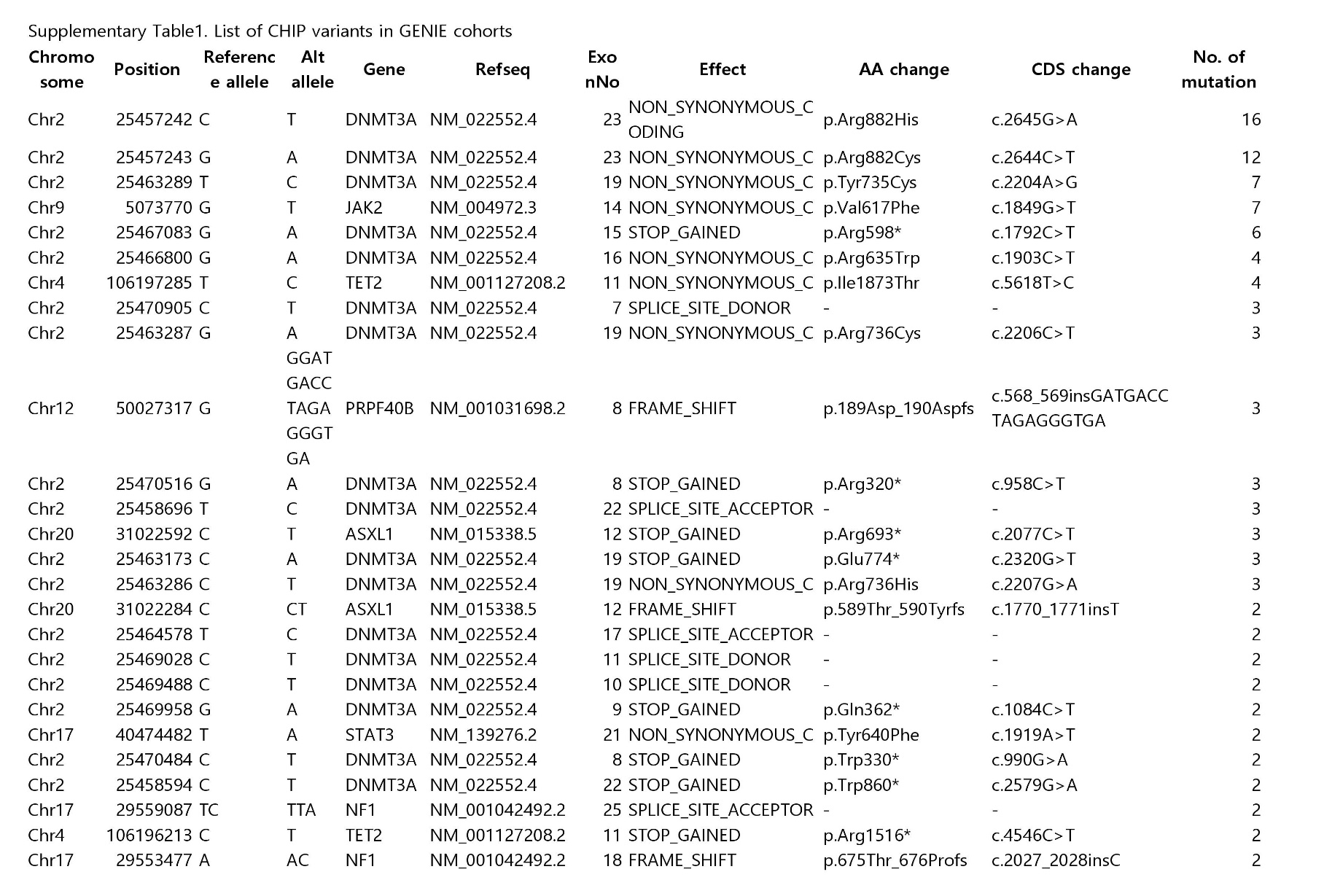

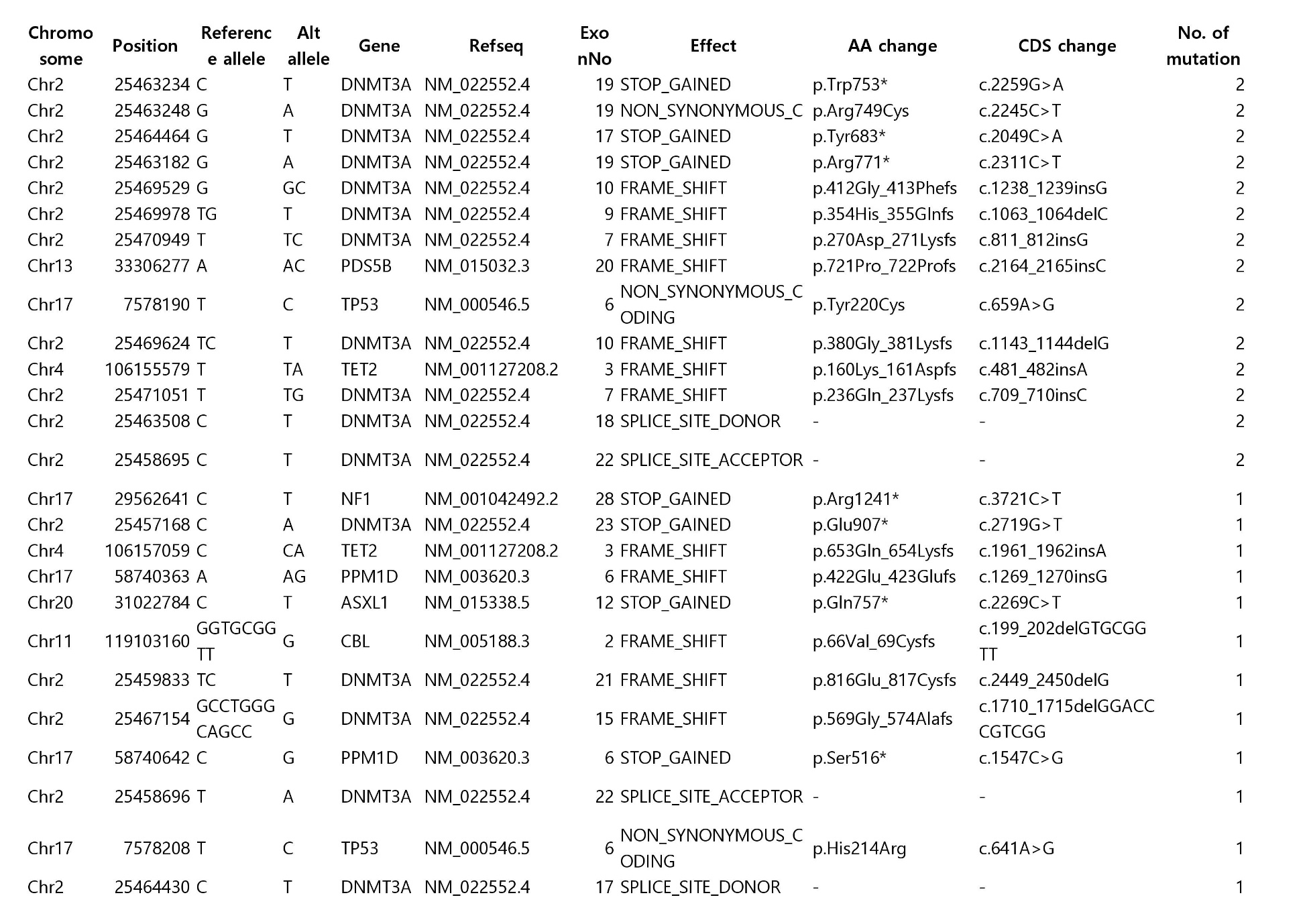

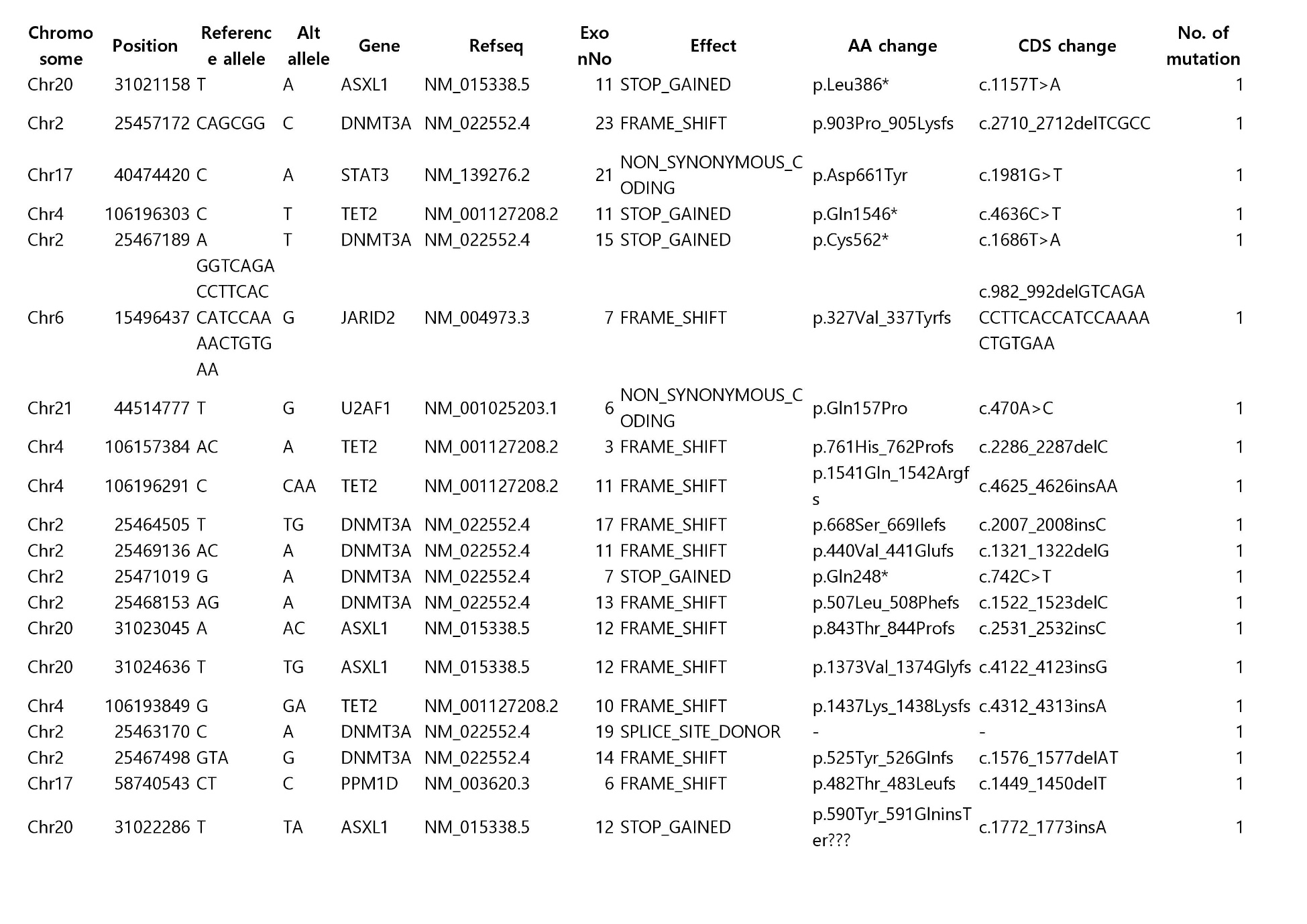

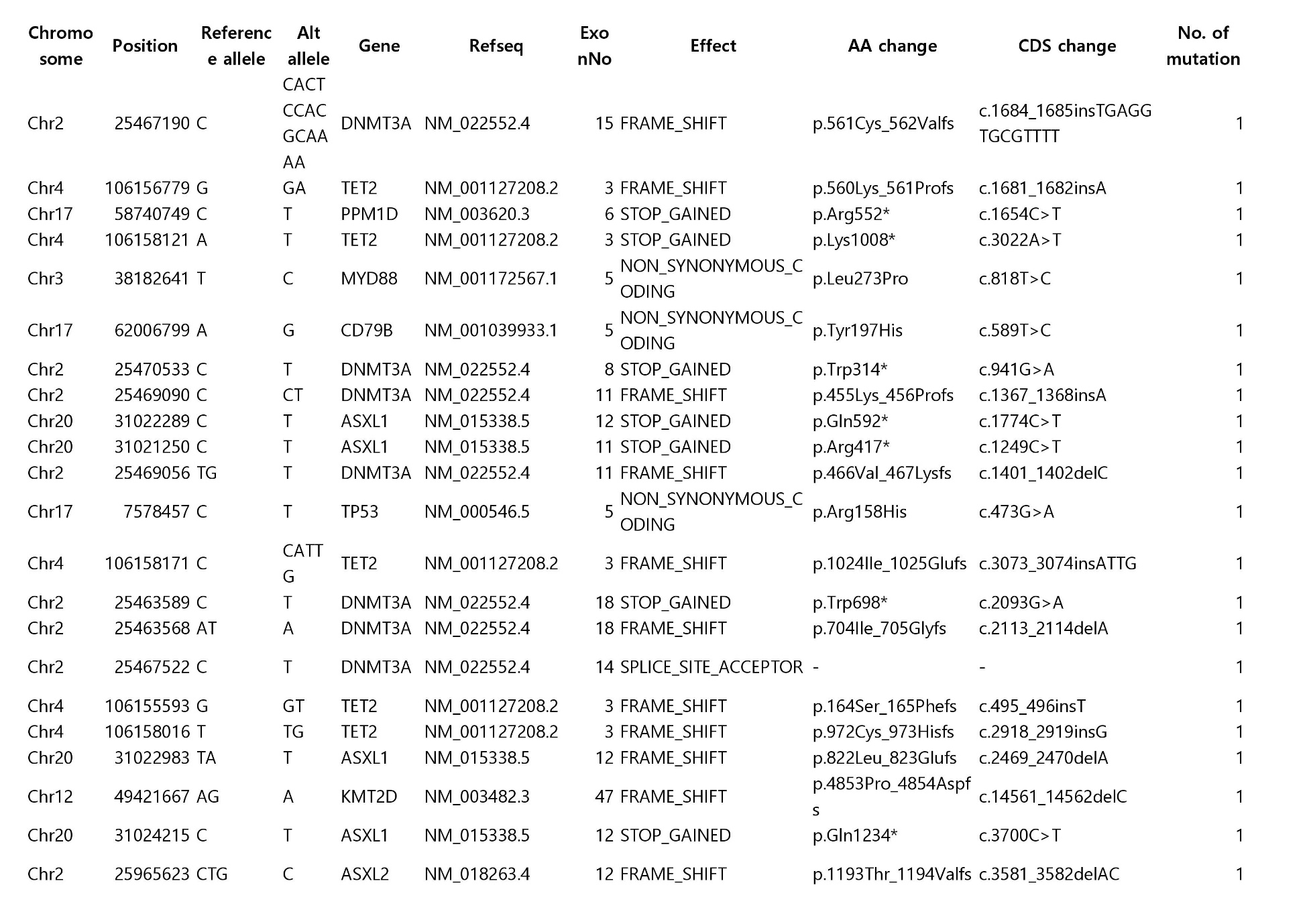

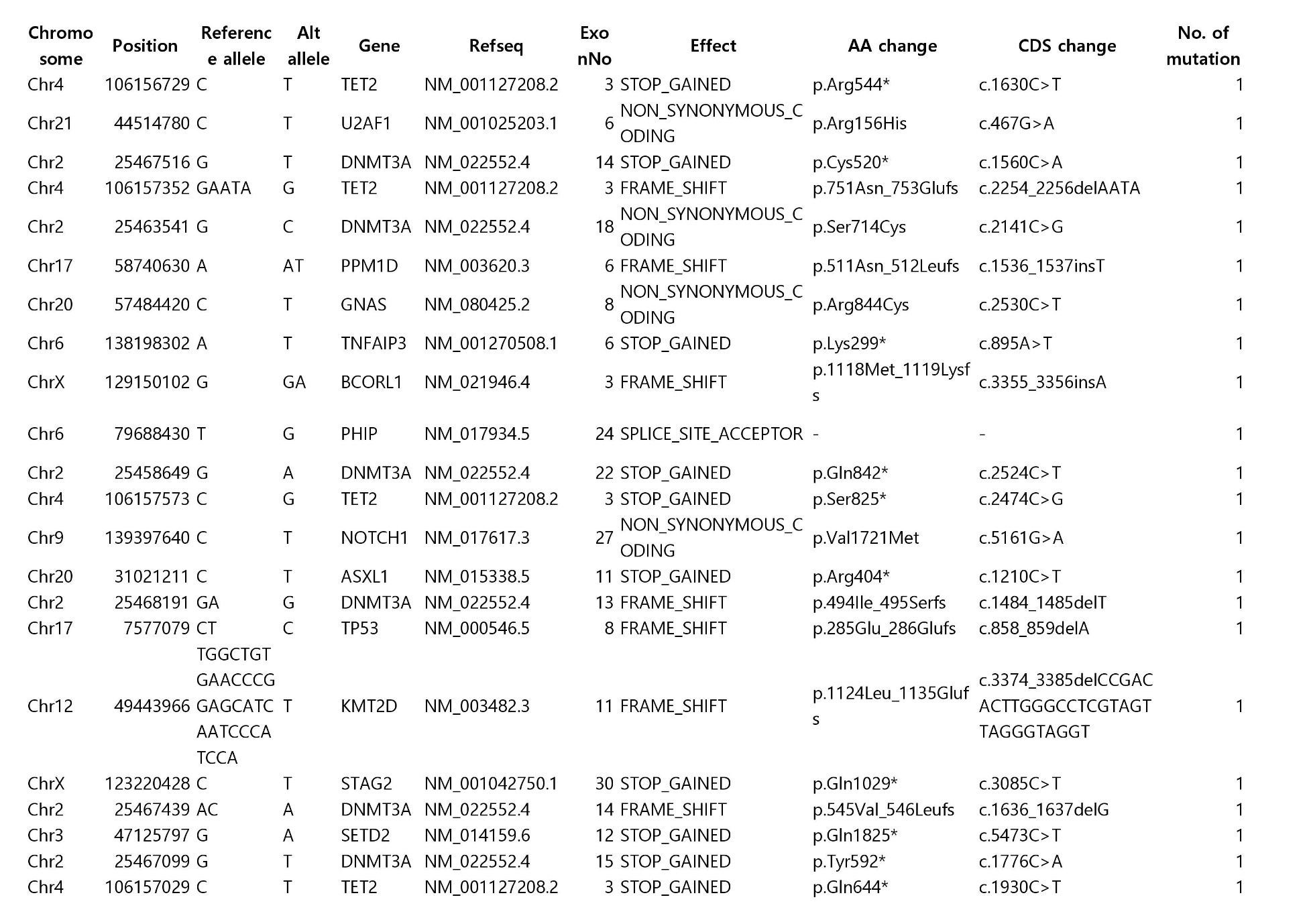

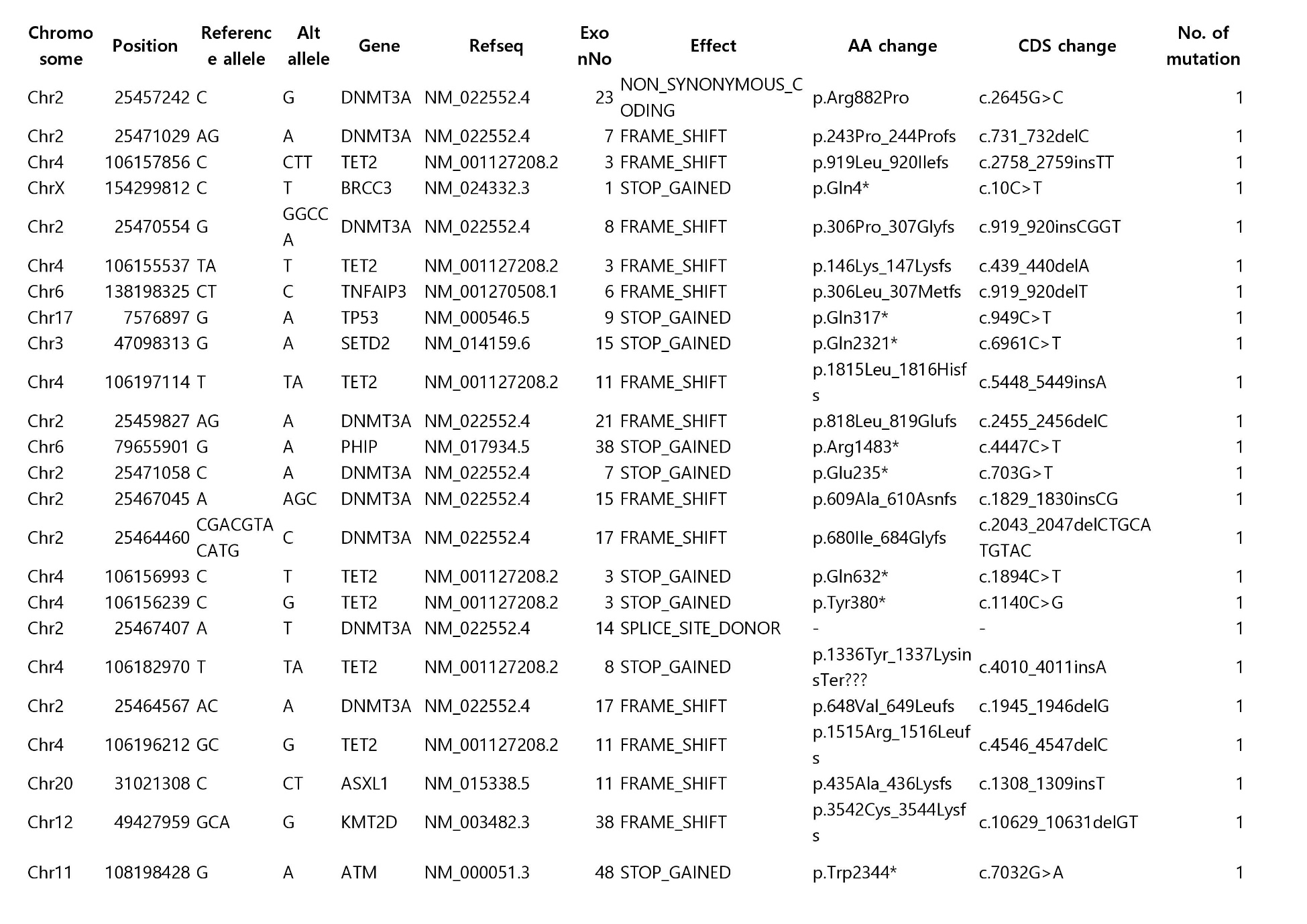

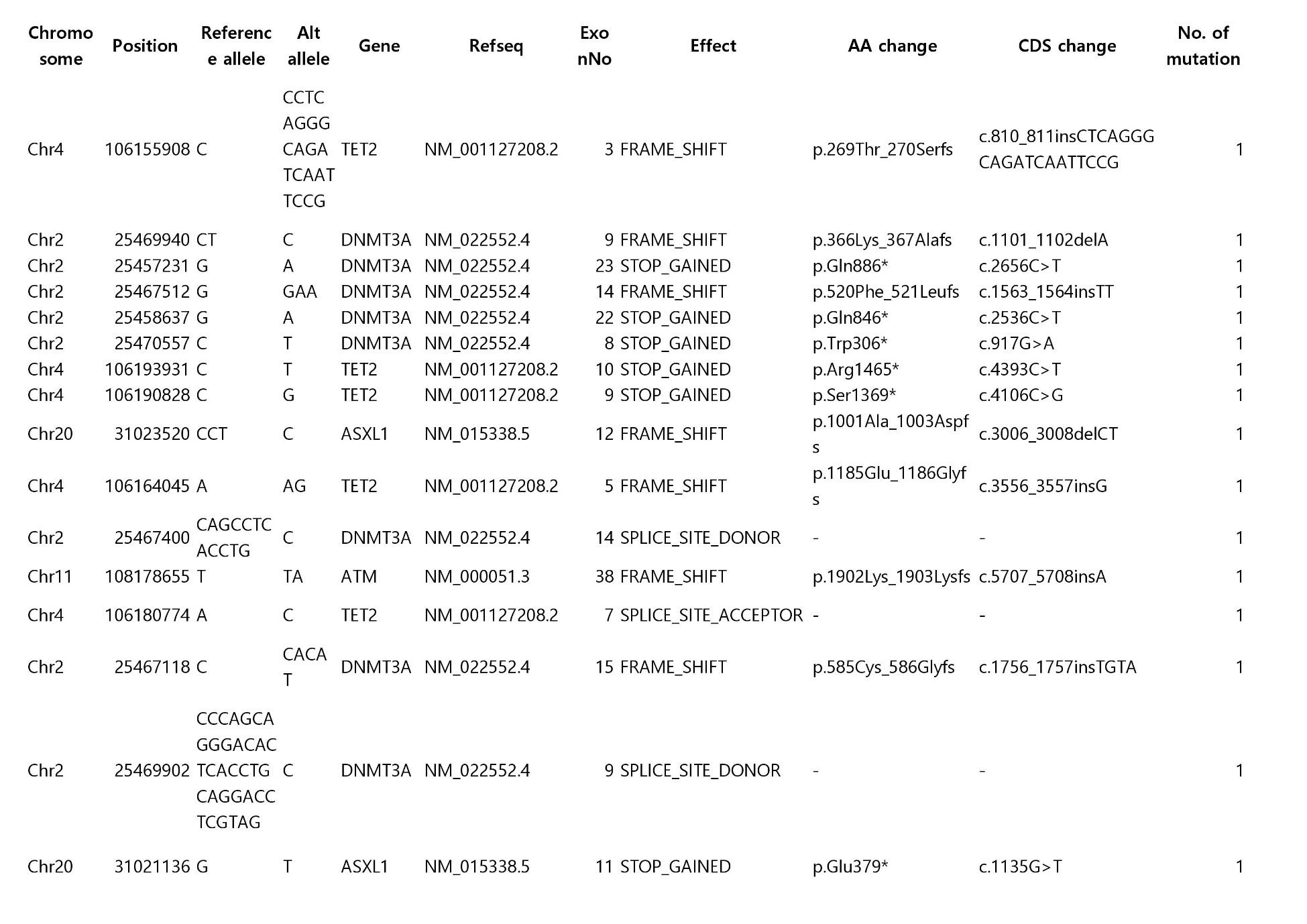

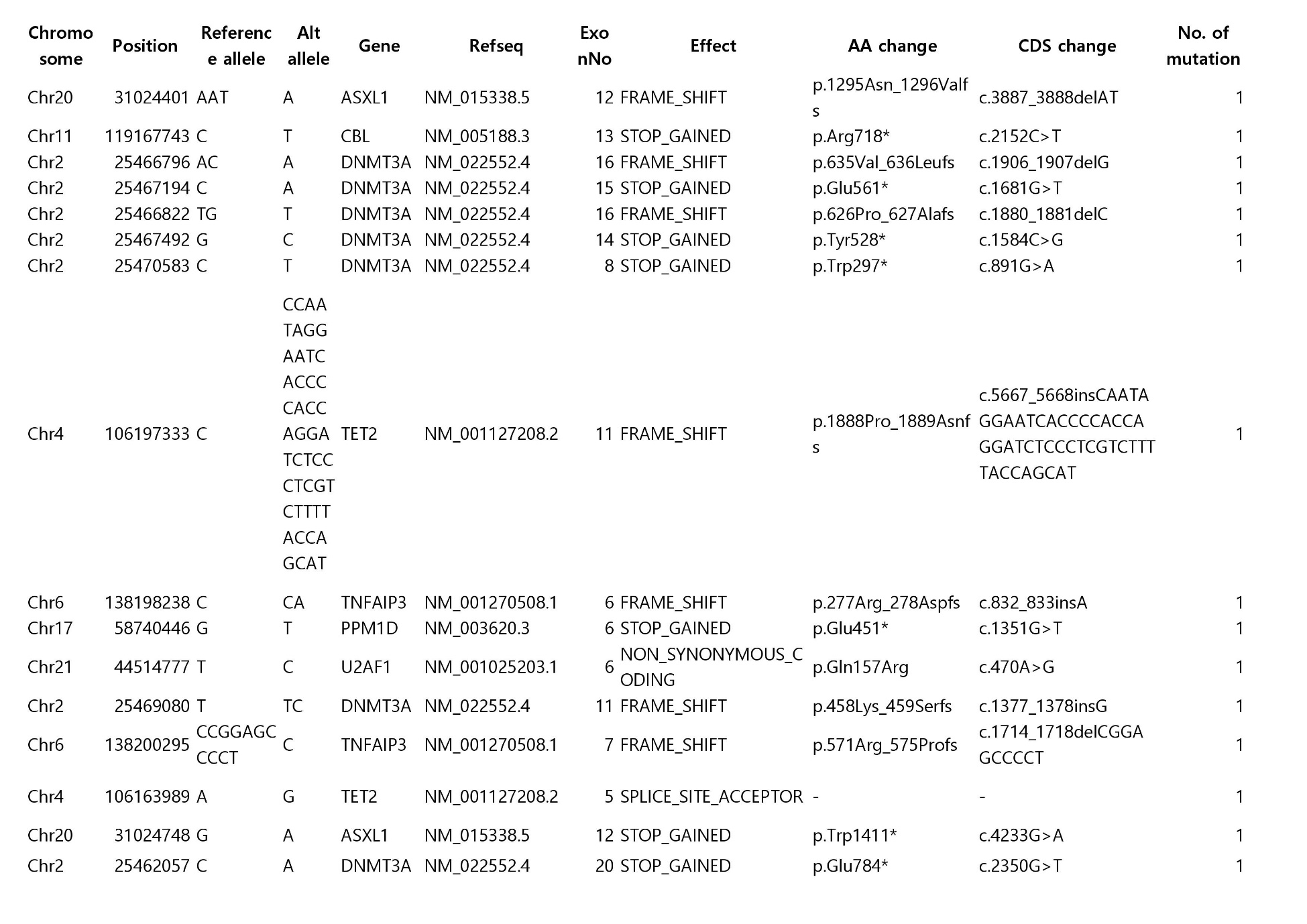

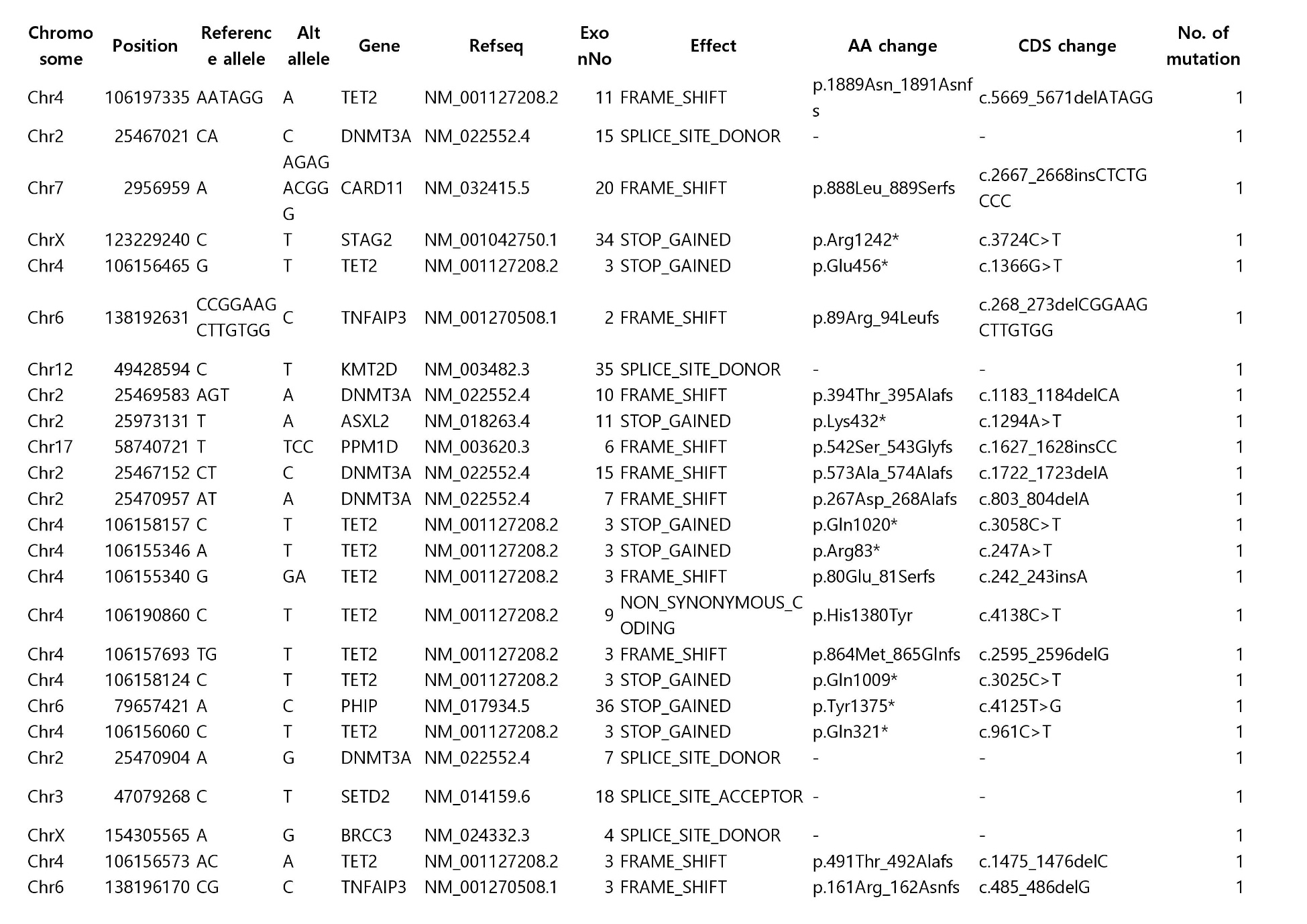

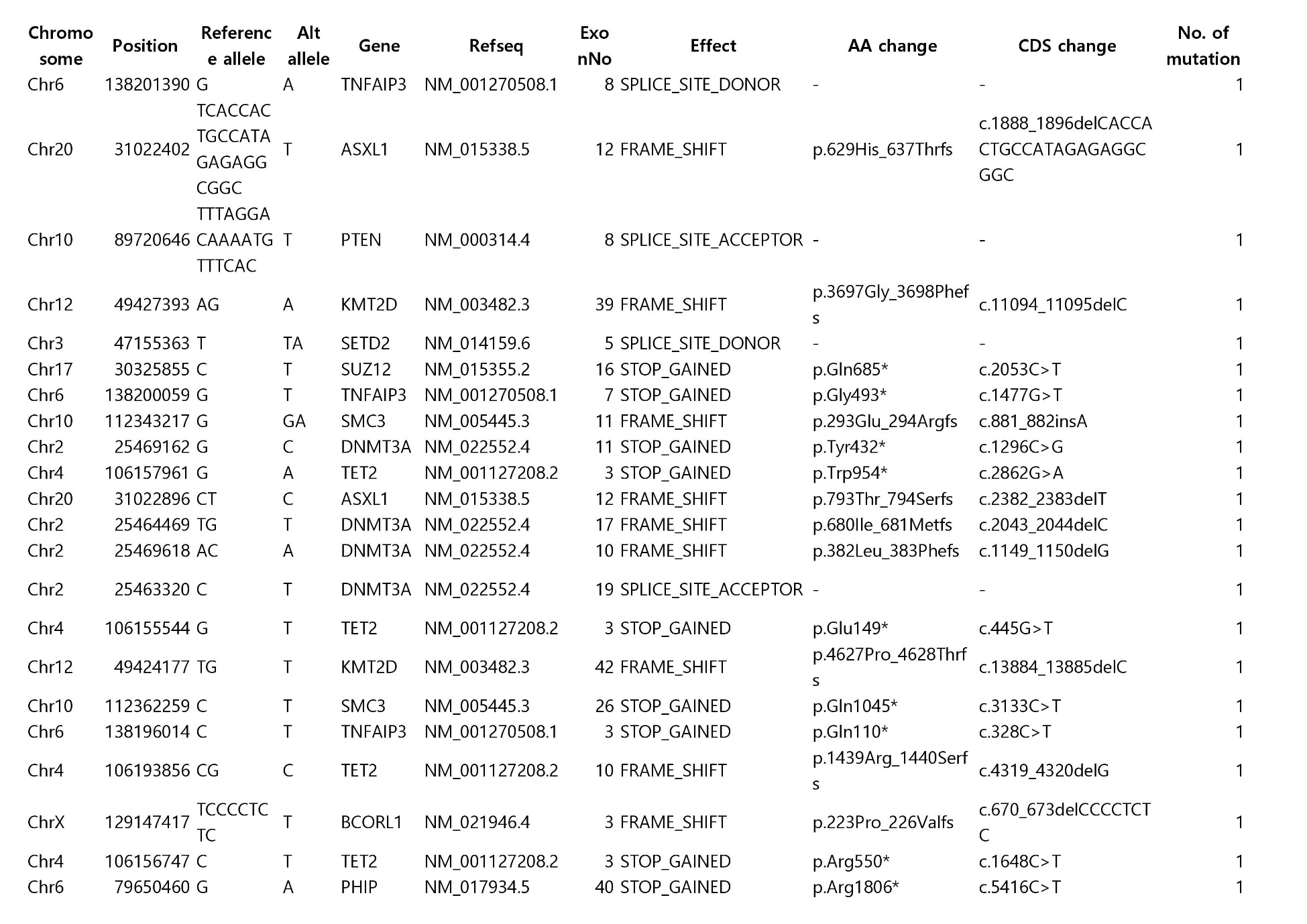

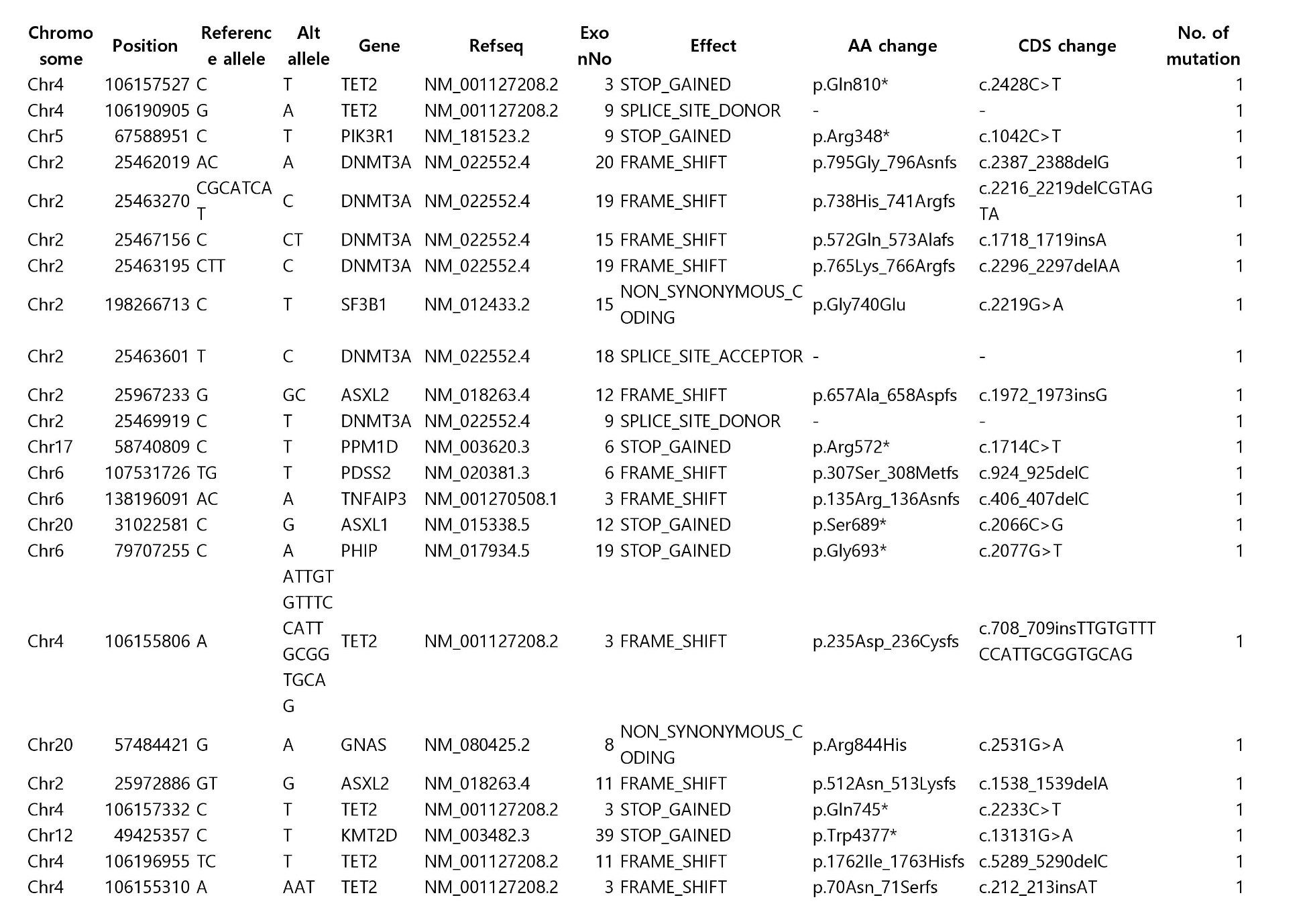

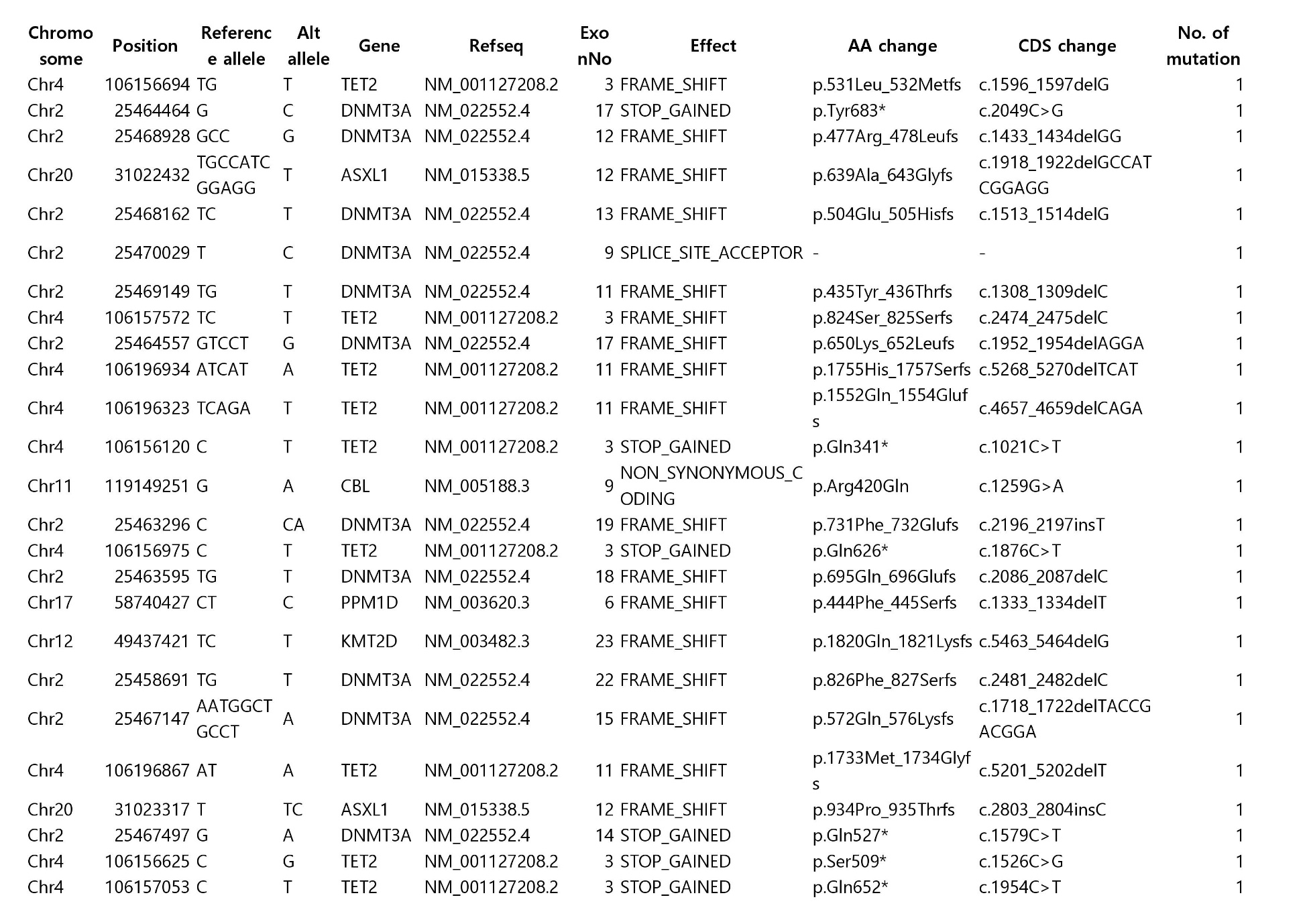

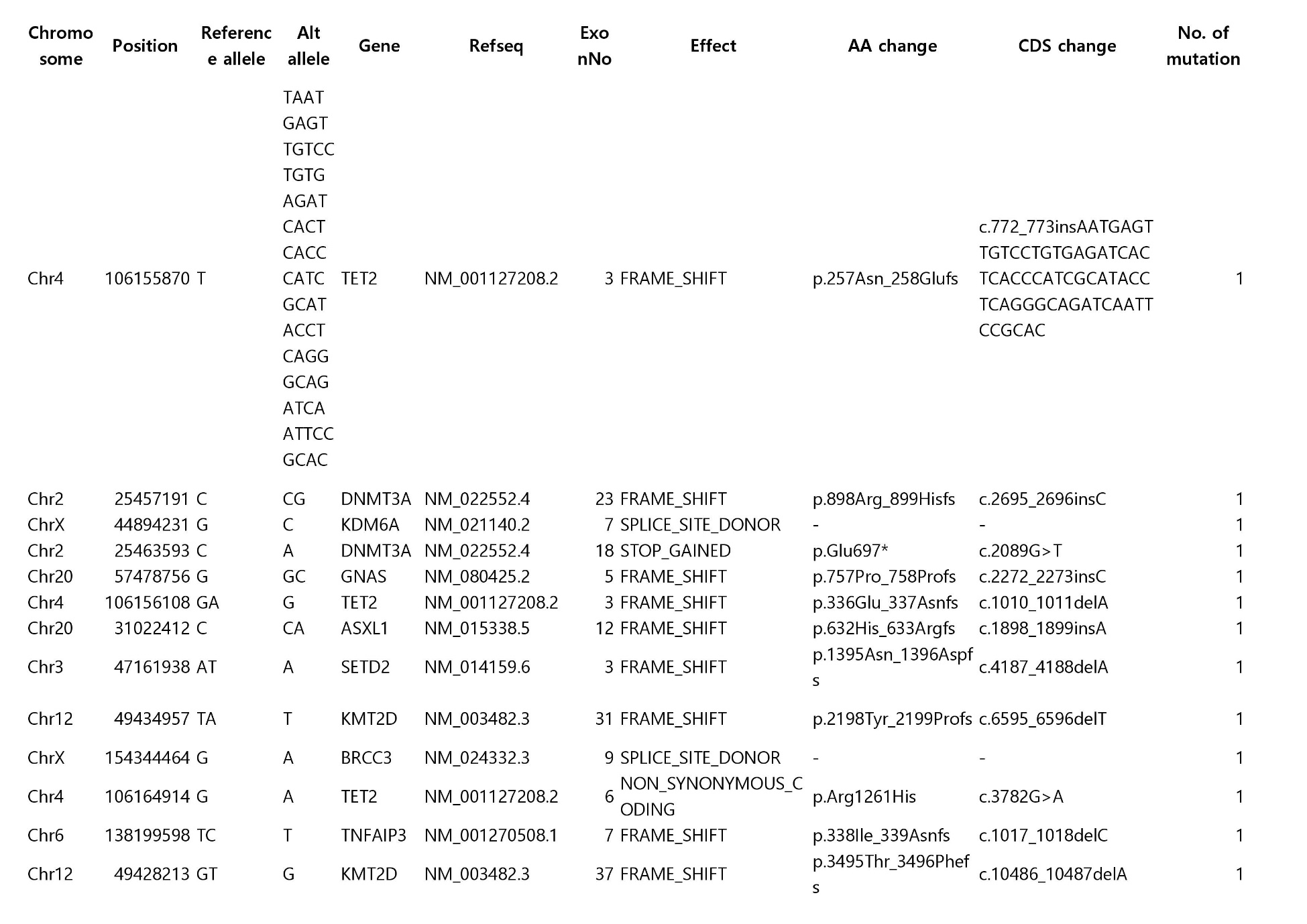

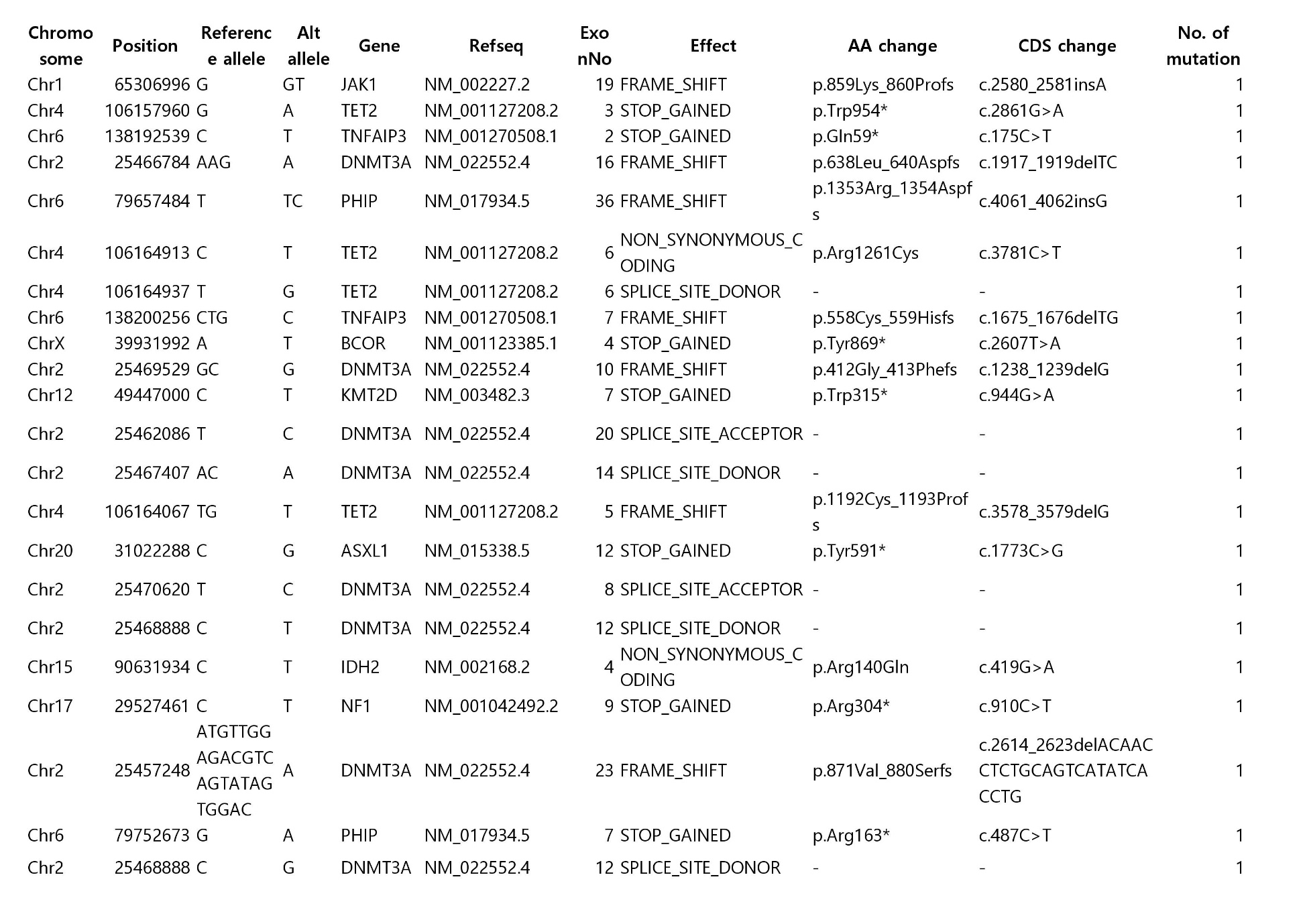

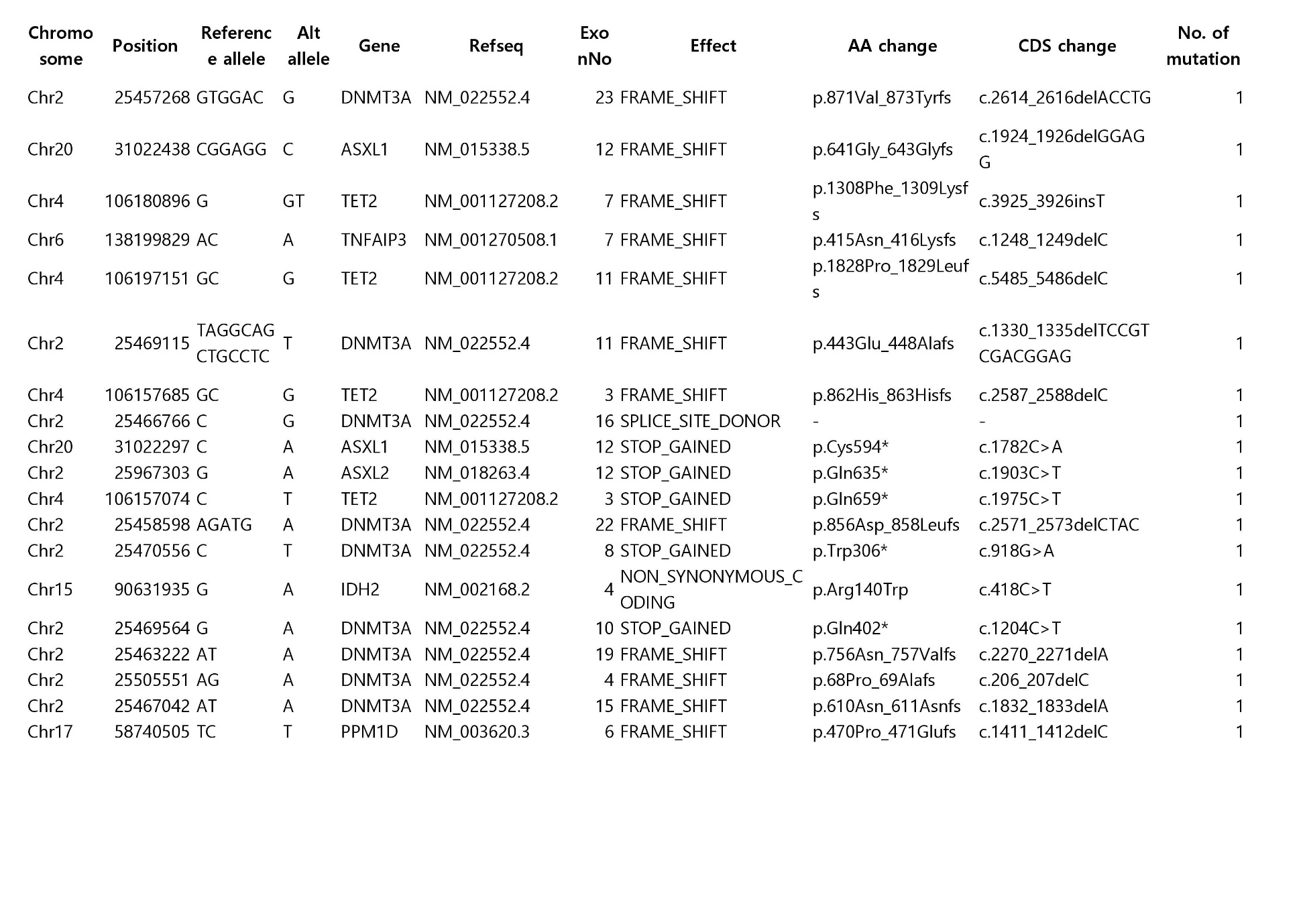

CHIP, clonal haematopoiesis of indeterminate potential

**Table S2. Incidence of clinical events according to CHIP status**

|  | Total  (n = 4,300) | CHIP carrier  (n = 368) | Non-carrier  (n = 3,932) |
| --- | --- | --- | --- |
| Cardiovascular death | 1 | 0 | 1 |
| Acute myocardial infarction | 2 | 0 | 2 |
| Revascularization^*^ | 56 | 9 | 47 |
| Definite angina  with medical therapy | 6 | 3 | 3 |
| Stroke | 12 | 5 | 7 |
| Carotid artery disease  with intervention | 0 | 0 | 0 |
| Aortic aneurysm  with intervention | 0 | 0 | 0 |
| Peripheral artery disease  with intervention | 3 | 1 | 2 |
| Follow-up (median, IQR) | 4.7 (3.2 – 5.3) | 4.9 (3.3 – 5.3) | 4.7 (3.1 – 5.3) |
| Total ASCVD | 80 | 18 | 62 |

**^*^**Revascularization includes percutaneous coronary intervention and coronary artery bypass graft. ASCVD, atherosclerotic cardiovascular disease; IQR, interquartile range; other abbreviations as **Table S1**.

**Table S3. Synergism between conventional risk factors and CHIP mutations**

|  | CHIP carrier | | Non-carrier | | Synergy index  (95% CI) | *p* |
| --- | --- | --- | --- | --- | --- | --- |
| **Risk factors** | **No** | **Yes** | **No** | **Yes** |  |  |
| Hypertension | 224 | 144 | 2,342 | 1,574 | 2.04 (0.60 – 6.93) | 0.256 |
| High total cholesterol | 256 | 112 | 2,704 | 1,227 | 2.66 (0.78 – 9.05) | 0.117 |
| High triglycerides | 318 | 49 | 3,338 | 580 | 0.62 (0.04 – 9.48) | 0.744 |
| High LDL cholesterol | 255 | 113 | 2,597 | 1,335 | 4.61 (1.04 – 20.57) | 0.045 |
| Low HDL cholesterol | 281 | 86 | 3,128 | 790 | 1.12 (0.15 – 8.10) | 0.920 |
| Diabetes mellitus | 312 | 52 | 3,423 | 490 | 3.00 (0.91 – 9.86) | 0.069 |
| Obesity | 234 | 130 | 2,435 | 1,461 | 1.24 (0.29 – 5.34) | 0.790 |
| Smoking | 291 | 77 | 2,825 | 1,107 | 1.14 (0.22 – 5.99) | 0.890 |

LDL, low-density lipoprotein; HDL, high-density lipoprotein; CI, confidence interval; other abbreviations as **Table S1**.

**Table S4. Imaging characteristics of baseline CCTA according to CHIP status**

|  | CHIP carrier  (n = 74) | Non-carrier  (n = 877) | *p* |
| --- | --- | --- | --- |
| ASCVD event | 5 (8.8) | 18 (2.6) | 0.024 |
| Age at baseline CCTA | 57.0±6.6 | 53.9±6.7 | <0.001 |
| Age at CHIP test | 61.1±7.4 | 57.2±7.3 | <0.001 |
| Time interval between CCTAs | 63.6(48.6-104.1) | 65.9(48.7-91.5) | 0.586 |
| Male | 66 (89.2) | 785 (89.5) | 0.845 |
| CACS | 19.5 (0.0-131.5) | 2.0 (0.0-52.0) | 0.212 |
| CACS 0 | 30 (40.5) | 414 (47.2) | 0.278 |
| CACS 1-99 | 19 (25.7) | 267 (30.4) | 0.431 |
| CACS 100-399 | 13 (17.6) | 111 (12.7) | 0.213 |
| CACS ≥400 | 8 (10.8) | 43 (4.9) | 0.052 |
| Presence of any plaque^*^ | 42 (56.8) | 479 (54.6) | 0.808 |
| Calcified plaque | 37 (50.0) | 372 (42.4) | 0.222 |
| Mixed or non-calcified plaque | 16 (21.6) | 220 (25.1) | 0.577 |
| Maximal focal diameter stenosis, % | 37.4±17.1 | 31.1±13.3 | 0.009 |
| Presence of plaque with diameter stenosis >50% | 6 (8.1) | 27 (3.1) | 0.037 |
| Number of segments with any plaque | 1.9±2.4 | 1.4±2.0 | 0.103 |
| Number of vessels with any plaque | 1.18±1.27 | 1.01±1.15 | 0.228 |
| Any involvement in LM or pLAD | 33 (44.6) | 347 (39.6) | 0.459 |
| Diameter stenosis >50% in LM or pLAD | 3 (4.1) | 11 (1.3) | 0.088 |

Values are mean ± standard deviation, median (interquartile range), or n (%). **^*^**Presence of plaques in at least one segment was denoted. CCTA, coronary computed tomography angiography; ASCVD, atherosclerotic cardiovascular disease; CACS, coronary artery calcium scores; LM, left main; pLAD, proximal left anterior descending coronary artery; other abbreviations as **Table S1**.
